# Expecting the unexpected: Subjective life expectancy and actual mortality

**DOI:** 10.64898/2026.08.24.26361278

**Authors:** Shohei Okamoto, Atsuhiro Yamada, Erika Kobayashi, Jersey Liang

## Abstract

**Objective:** This study evaluated how well subjective life expectancy (SLE) predicts mortality and actual life expectancy (ALE), along with factors associated with inaccurate expectations.

**Methods:** Using panel data on approximately 2,000 individuals with up to 28 years of follow-up from a nationally representative sample of older Japanese adults, we examined relationships among SLE, actual mortality, and ALE by survival analysis. We also evaluated health and socioeconomic disparities using concentration indices and investigated factors influencing SLE–ALE discrepancies and focal-point (i.e. rounded or anchored estimates) and do-not-know responses. SLE was measured as a self-reported point estimate, whereas ALE mainly came from official records and family reports.

**Results:** SLE was significantly associated with both actual mortality and ALE, even after accounting for demographic and socioeconomic variables. Nonetheless, significant inaccuracies remain: approximately 59% of individuals surpassed their expected lifespan. SLE was positively associated with ALE; however, the association was inelastic. Women and those with higher education levels were more likely to outlive their SLE, whereas those in poorer health were less likely to do so. Higher education correlated with fewer focal point responses to the SLE question.

**Discussion:** SLE effectively predicts ALE; however, gaps are non-negligible and differ across gender and socioeconomic groups. Offering more precise data, such as sex- and age-specific remaining life expectancy, can enhance SLE formation and lead to more informed economic choices.

**Highlights:**

- Subjective life expectancy is linked to actual mortality and life expectancy.
- Significant inaccuracies in subjective life expectancy remain.
- Approximately 59% of individuals outlived their expected lifespan.
- Women and the better-educated were more likely to outlive their expectations.
- Providing sex- and age-specific data can improve one’s lifespan expectations.

## 1. Introduction

### 1.1. Background

Longevity allows individuals to enjoy longer lives but generates uncertainties about preparedness for an extended lifespan. Universal old-age pension schemes, common in many developed countries, help smooth consumption throughout life, especially after retirement. Nevertheless, population ageing and declining fertility threaten the sustainability of these programmes. Consequently, numerous nations are reforming pension and labour policies, such as encouraging later retirement by offering financial (dis)incentives and adjusting coverage, contribution rates, and benefit levels (OECD, 2025). When introducing greater flexibility and less generous benefits to maintain financial balance, these reforms shift greater responsibility for life planning, including retirement, annuity, and lifetime wealth planning, to individuals. This life-planning challenge is often addressed through financial literacy to improve post-retirement decision-making (Lusardi & Mitchell, 2014). Nonetheless, uncertainty about lifespan complicates planning since people lack precise knowledge of how long they will live, known as subjective life expectancy (SLE).

Japan is at the forefront of population ageing, with the life expectancy at birth reaching 81.1 years for men and 87.1 years for women in 2024, while remaining life expectancy at age 65 being 19.5 and 24.3 years, respectively (Ministry of Health, Labour and Welfare, 2025). Similar to other high-income countries, the old-age dependency ratio, the proportion of people aged 65 or older relative to the working-age population aged 15–64, is expected to rise from 0.50 in 2026 to 0.70 in 2050 (National Institute of Population and Social Security Research, 2023). This presents challenges to sustainable social policy financing, including the pay-as-you-go pension system. As the workforce shrinks and longevity increases, pension benefits are expected to decline, requiring individuals to make up for reduced retirement income. Consequently, the Japanese government has promoted asset accumulation among citizens through instruments such as tax-exempt investment accounts (i.e. the Nippon Individual Savings Account) and defined contribution plans, increasing personal responsibility for financial management. However, financial literacy levels in Japan are low (Sticha & Sekita, 2023), which hampers wealth accumulation, while retirement wealth decumulation also remains suboptimal, mainly due to precautionary saving behaviours (Niimi & Horioka, 2019). Longer lifespans than in other countries may also heighten uncertainty regarding lifespan; thus, understanding SLE among the population is crucial for helping individuals plan their financial and economic behaviours in line with their longevity expectations.

### 1.2. *Predictive validity, determinants, and consequences of SLE*

The life-cycle hypothesis posits that individuals optimise their consumption and saving decisions throughout their lives. Despite its centrality to the theory, no attention had been paid to SLE until the study by Hamermesh (1985). Since then, SLE have been researched to assess its association with actual mortality and mortality tables, determinants, and economic behaviours as described below.

#### (1) Predictive validity and formation of SLE

SLE’s predictive validity was evaluated by comparing it with actual mortality outcomes and mortality tables in the United States (Bago d’Uva et al., 2020; Foltyn & Olsson, 2024; Hurd & McGarry, 2002; Smith et al., 2001), England, Ireland, and Scotland (Bell et al., 2020), the Netherlands (van Solinge & Henkens, 2018), a larger number of European countries (Post & Hanewald, 2013), and the Republic of Korea (Bae et al., 2017; Kim & Kim, 2017). Although these studies identified positive associations between SLE and actual mortality and actuarial estimates, gaps exist (Bago d’Uva et al., 2020), showing variability across demographic, socioeconomic, and health-related characteristics.

The way SLE is formed can contribute to these gaps. People may base their lifespan expectations on private knowledge, including their own health status, which significantly influences actual life expectancy (ALE) (B. Griffin et al., 2013), along with various reference points. These include the average life expectancy at birth and average remaining life expectancy at their current age, both influenced by demographic characteristics (e.g. age and gender). Although age- and sex-specific average remaining life expectancy is a more appropriate benchmark, many may rely on the more familiar overall average at birth, leading to a wider SLE–ALE gap by expecting shorter lives than actuarial estimates. The ages at death of others (e.g. parents, partners, and close friends) can also serve as reference points (Donnelly et al., 2020), influencing individuals’ attitudes towards death and their expectations about its timing. Limited cognitive ability and numeracy induce bounded rationality, impacting how well individuals interpret these information sources when estimating SLE and making decisions, as evidenced by the least educated tending to be the least accurate in their predictions (Bago d’Uva et al., 2020).

Another reason for the gaps is focal point responses, including rounding (e.g. to multiples of 5 or 10 years) and anchoring (e.g. responses of 80 or 85 years based on the salient overall average life expectancy at birth). Estimating one’s own life expectancy involves uncertainty, which leads to specific reporting behaviours, including focal-point responses (Ruud et al., 2025) and do-not-know answers. These behaviours are common in SLE reporting and can cause discrepancies from ALE (Kleinjans & Soest, 2013; Rappange et al., 2017). However, little is known about who is most likely to engage in these reporting behaviours. Understanding this could help identify individuals who may need extra assistance with financial and life planning based on their SLE.

#### (2) Health and economic behaviours

Based on the classical human capital theory (Grossman, 2000), an individual’s lifespan is considered endogenous, influenced by utility maximisation involving investments in health stock that deteriorates with age over their lifespan. This framework explains the links among health, health behaviours, and SLE, aligning with prior research indicating that unhealthier lifestyles and current poor health are associated with lower SLE and less optimistic health outlooks for the future (Bae et al., 2017; Li et al., 2024; Rappange et al., 2016a, b).

A key area of focus in the literature is how SLE impacts economic behaviours, including retirement and financial decisions. According to the life-cycle hypothesis, rational individuals aim to smooth consumption throughout their lives; thus, those with longer SLE are expected to delay retirement and pension claims. Socioemotional selectivity theory, which explains motivation across the lifespan, suggests that perceived remaining lifetime influences behaviour—either towards gaining knowledge and experience for the future or managing emotional states to enhance well-being (Carstensen, 2006). When individuals anticipate a longer life, they see death as distant and feel they have enough time for work and leisure, reducing the motivation to retire early. They might stay in the workforce longer to build financial resources for a longer retirement period. Barbara Griffin et al. (2012) found that SLE predicts intended retirement age, financial and non-financial preparation, and actual retirement over a year. van Solinge and Henkens (2010) found that longer SLE correlates with later retirement intentions, though these intentions are not always realised within 5–6 years. Conversely, in the United States, those with very low SLE tend to retire earlier than those expecting to live longer (Hurd et al., 2004). Similarly, Yang et al. (2024) reported that SLE positively impacts labour supply in China, with rural-urban heterogeneity affecting both extensive and intensive margins (i.e. labour participation and workhours).

Linked to retirement behaviours, SLE also affects decisions about savings and claiming old-age pensions. Those with poorer health often underestimate their life expectancy, shaping their time preferences and saving habits, which contributes to health-related wealth gaps (Foltyn & Olsson, 2024). Nivalainen (2022) showed that individuals factor their expected longevity into pension claiming decisions, with shorter expectations prompting earlier claims. Other research has also shown a positive link between SLE and early Social Security benefit claims, though these effects are small and SLE alone cannot explain claiming behaviour (Dai et al., 2023; Hurd et al., 2004). Brinch, Fredriksen, & Vestad (2018) indicated that predicted longevity, based on observable traits, influenced old-age pension claims, thereby boosting individuals’ financial benefits, though the added costs to public funds were modest.

### 1.3. *SLE measurement*

To elicit SLE, subjective probabilities and point estimates are used (Rappange et al., 2017). The first method requires individuals to state the percentage chance they believe they will live to specific ages. Economics generally interprets decision-making as aimed at maximising expected lifetime utility based on observed behaviour, and nowadays, such expectations are increasingly gauged through subjective probabilities (Manski, 2004). While this method can be more accurate than point estimates, since death is inherently stochastic, it has been adopted in surveys like the Health and Retirement Study (Foltyn & Olsson, 2024) and the Survey of Health, Ageing and Retirement in Europe (Rappange et al., 2016b). However, it is cognitively demanding, often leads to inconsistent answers, and does not directly measure the individual’s subjective expected lifespan.

In contrast, the second approach asks individuals for a single, non-probabilistic estimate of their expected lifespan or time of death. While useful for directly estimating SLE, it does not capture uncertainty about the timing of death. Compared to the subjective probability approach, it is more respondent-friendly due to its simplicity and clarity. Although it has been used in several cross-sectional surveys (Brouwer & van Exel, 2005; Rappange et al., 2017), it has not yet been applied to longitudinal data from a population-based representative sample, so there is limited evidence on how accurately it predicts individual longevity.

Both methods frequently feature focal point responses (Rappange et al., 2017), highlighting the importance of additional research to determine which individuals tend to show these behaviours. This can aid in more accurately forming SLE and improve future decision-making. Additionally, more insights are needed on the point-estimate approach, which is less demanding for respondents, to evaluate its effectiveness in predicting actual mortality and longevity.

### 1.4. *Objectives and contributions of this study*

Given the limited evidence on SLE, especially regarding the point-estimate approach, which is less cognitively demanding, directly comparable to actual lifespan, yet potentially less precise than the subjective probability approach, this study aims to evaluate the prognostic value of SLE as measured by the point-estimate technique. The study addresses this evidence gap in four ways. First, analysing data from a nationally representative sample of older Japanese adults with up to 28 years of follow-up enables us to compare SLE with actual ages at death for nearly 1,500 individuals. This study therefore provides valuable insights into the predictive validity of SLE using an exceptionally well-suited dataset. Second, it enhances understanding of the factors influencing SLE and the gap between expected and actual lifespan, by examining socioeconomic and health-related disparities in SLE accuracy. Third, it explores the factors affecting focal point responses to the SLE question to identify contributors to potential inaccuracies in SLE estimation. Fourth, this study provides evidence on SLE from Japan, a context that has received little attention in the existing literature. Given that Japan has one of the longest life expectancies in the world, the findings contribute evidence from a unique context and offer important policy-relevant implications.

## 2. **Methods**

### 2.1. Data

This study examined data from the National Survey of the Japanese Elderly (NSJE), a nationally representative sample of individuals aged 60 and above in Japan. Participants were selected using a two-stage stratified random sampling method that considered regions and population sizes. The initial survey (Wave 1) took place in 1987, with subsequent follow-ups every 3–6 years that included new samples. The latest survey was conducted in 2024 (Wave 11).

This study mainly analysed data from Wave 4 (1996), which included a question about SLE. Wave 4 involved follow-up respondents from cohorts recruited in Wave 1 (1987) and Wave 2 (1990), along with a newly added sample. Additional details of the survey are available elsewhere (JAHEAD/NSJE Project Group, n.d.). The Tokyo Metropolitan Institute for Geriatrics and Gerontology’s ethical review committee approved the study (No. R24-033).

### 2.2. Subjective and actual life expectancies

The Wave 4 questionnaire included a variable measuring SLE, asking respondents the age they expect to reach. Responses ranged from 63 to 120 years, with 393 out of 2,447 respondents indicating that they did not know. Compared to those who provided specific ages, this group had a higher proportion of women (62.8% vs. 55.2%), was slightly older on average (71.8 vs. 69.7 years), and reported poorer health more often (20.0% vs. 11.6%).

Of the 2,054 respondents who provided valid responses to the SLE question, 1,514 passed away over 28 years from Wave 4 (post-survey) to Wave 11. In nearly all cases (approximately 98.4%), death dates were sourced from resident records or family reports. When the death date was missing, we estimated it by assuming the individual died halfway between the survey waves when the death was recorded.

To evaluate the accuracy of SLE, we calculated the ratio of ALE to SLE, using it as either a continuous or dummy variable. When treated as a continuous variable, the ratio indicates (1) shorter-than-expected survival when it is less than one and (2) longer-than-expected survival when it is greater than one. The dummy variable includes four categories: (1) Equal, representing individuals whose SLE corresponded to their ALE; (2) Long-lived, indicating individuals whose ALE exceeded their SLE, as well as those who were still alive in Wave 11 and had already exceeded their SLE; (3) Short-lived, denoting individuals whose ALE was shorter than their SLE; and (4) DK, indicating individuals who reported that they did not know their SLE. Individuals who were still alive and had not yet reached their SLE were excluded from this variable because it was not yet possible to determine whether their ALE would exceed, match, or fall short of their SLE.

### 2.3. Empirical strategies

We conducted three analyses to assess how accurately individuals predict their longevity and the determinants of prediction accuracy.

#### Predictability of SLE for actual mortality

We initially assessed whether SLE could reliably predict mortality by analysing data from Wave 4 respondents, including both survivors and decedents from Wave 4 to Wave 11. A parametric survival analysis examined how subjective remaining life expectancy related to actual survival over a follow-up period of up to 28 years. A Gompertz distribution was selected based on the Akaike Information Criterion, compared against exponential, log-logistic, Weibull, log-normal, and generalised gamma models. From the analysis, we estimated predicted marginal survival times by subjective remaining life expectancy to assess the correspondence between SLE and ALE. To facilitate visualisation, the SLE variable was also transformed into quartiles to compare survival curves across its different levels.

#### Socioeconomic and health disparities

We examined SLE-ALE discrepancies across socioeconomic and health statuses, by calculating the concentration index (CI) (O’Donnell et al., 2008; Wagstaff, 2005). The CI measures the area between the concentration curve—which shows the proportion of SLE-related indicators explained by the cumulative share of individuals ordered from lowest to highest in the variable—and the 45-degree line representing perfect equality. The CI ranges from –1 to 1: negative values indicate concentration among those with lower education or poorer health, while positive values indicate concentration among those with higher education or better health. For instance, a negative CI for the ratio of ALE to SLE, using health as the rank variable, suggests that individuals with poorer health tend to live longer lives than expected (i.e. SLE ≤ ALE), with the concentration curve lying above the 45-degree line.

#### Determinants

We performed regression analyses to investigate factors influencing SLE and its variation from ALE. The analysis used remaining SLE as the continuous dependent variable and the categorical indicator denoting whether SLE was equal to, greater than, or less than ALE, or was reported as unknown. A linear model was applied to the continuous measure, while a multinomial logistic regression model was used for the categorical outcome.

Focal point responses can lead to discrepancies between ALE and SLE. To better understand what influences these reporting behaviours, we examined the factors behind focal point responses to the SLE question. We classified focal point responses as those given at five- or ten-year intervals, as well as at age 80, which approximates the sex-aggregated life expectancy at birth. Additionally, we analysed responses for body height and weight to see if similar focal point patterns appear for questions that involve less uncertainty than life expectancy.

#### Independent variables

We used the following independent variables measured at Wave 4 and treated as time-constant to evaluate disparities and determinants. Since SLE is a self-reported, subjective measure, we included objective covariates to reduce common method variance.

#### Education

Cognitive and non-cognitive skills are connected to educational attainment (Malanchini et al., 2020). Together with numeracy, these skills impact the formation of SLE by affecting individuals’ ability to process pertinent information. We used respondents’ years of education as an indicator of these abilities to evaluate socioeconomic disparities and their underlying causes.

#### Health

Health is one of the major factors influencing SLE. Self-rated health, an overall assessment of one’s health status, is a well-known predictor of mortality (DeSalvo et al., 2006). However, there is a concern about common method bias arising from the self-reported nature of SLE. While objectively measured health can reduce this bias, such measures often lack the breadth to capture overall health comprehensively. To overcome this, we developed health scores by combining self-rated health with detailed health data (Bound et al., 1999; Coe & Zamarro, 2011), including physical and psychological health and health behaviours. Recognising that mental health partly overlaps with self-rated scales and that health behaviours may not fully reflect health status, we estimated four health scores (1) combining physical and mental health and behaviours, (2) with physical and mental health only, (3) with physical health alone, and (4) the original self-rated health. These scores were derived from the full NSJE dataset, which includes 20,400 observations from 6,823 individuals across Waves 1–11. Appendix Table A-1 lists the health-related variables and their descriptive statistics, while estimation results are available in Appendix Table A-2 and Appendix Figure A-1. Since the findings were similar for all four variables, we report only the results using the score (1) combining physical and mental health and health-related behaviours in most analyses.

#### Demographics

We included respondents’ age, age squared, gender, and marital status, differentiating between being single and bereaved. This distinction is important because spousal bereavement may affect SLE, with the spouse’s age at death acting as a reference point.

#### Socioeconomic status

To capture individuals’ socioeconomic status, we included employment status (whether currently employed or not) and house ownership, in addition to education.

#### Death of a close person

Experiencing the loss of a close person and the age at which they died can influence SLE formation by acting as reference points. While this specific detail was not available in the NSJE, the survey inquired whether participants had lost a family member or close friend in the past year. Dummy variables were created for each to partly account for the impact of these reference points.

#### Other control variables

We also included two variables to measure reference-point effects, based on official data from the Abridged Life Tables for Japan. The first is a dummy variable indicating if a respondent had already surpassed the sex-specific average life expectancy at birth in Wave 4 (77.0 years for men and 83.6 years for women). The second variable reflects the average remaining life expectancy at the respondent’s age and sex during Wave 4.

#### Sample size calculation

The sampling flow chart is shown in Appendix Figure A-2. The analysis was restricted to respondents who completed the Wave 4 survey independently and was conducted using three analytical samples: (1) those alive or deceased by the latest Wave 11 survey, (2) those who had died by Wave 11, and (3) individuals who had died by Wave 11, those who were still alive and had already exceeded their SLE, and those who reported “do not know” in response to the SLE question. Additionally, respondents with missing independent variable data were excluded, representing a small portion. However, as described below, multiple imputation was implemented as a robustness check to assess the sensitivity of the findings to missing data. The final sample sizes varied across models, ranging from 1,990 to 2,003 for sample category (1), 1,514 for sample category (2), and 1,820 for sample category (3).

#### Approach to sample attrition

Loss to follow-up occurs inevitably in sample surveys. Wave 4 respondents included both individuals newly recruited in Wave 4 and follow-up respondents from Waves 1 and 2. Response rates in Waves 1–4 ranged from 73.3% to 92.6% (see Appendix Table A-3). While the NSJE maintains relatively high response rates, there is still a concern about potential attrition bias.

To partly address this issue, we used inverse probability weighting (Seaman & White, 2013), combining cross-sectional and longitudinal weights. Cross-sectional weights were calculated as the inverse likelihood of response at baseline, based on participants’ age, gender, residential area, and municipality population category. Longitudinal weights represented the inverse probability of responding in each subsequent wave, conditioned on age, gender, marital status, education, employment, self-rated health, residential area, and municipality category at baseline. Additionally, our survival analysis examining the link between SLE and mortality minimised this concern, as it explicitly accounted for censoring and sample attrition.

#### Multiple imputation for item missingness

We observed missing values in several independent variables. To assess the robustness of the findings, we additionally conducted analyses using multiple imputation under the missing-at-random assumption. Information collected in Wave 4, including demographic, socioeconomic, psychological, and health-related variables, was used in the imputation model. Missing values were imputed using chained equations with predictive mean matching, generating 20 imputed datasets. We confirmed that the main findings remained unchanged after employing multiple imputation. Therefore, we interpret the results based on the complete-case analysis.

## 3. **Results**

### 3.1. Descriptive statistics

Table 1 presents descriptive statistics for the Wave 4 respondents, using primarily Wave 4 variables. Appendix Table A-4 also outlines the differences between alive and deceased individuals. The average age was approximately 70 years, with a slightly higher proportion of women. Compared to the deceased, the alive group was generally younger, had more women, longer educational histories, and better health scores. Additionally, subjective remaining life expectancy was higher among the alive respondents (16.3 years) compared to the deceased (12.0 years).

**Table 1.**
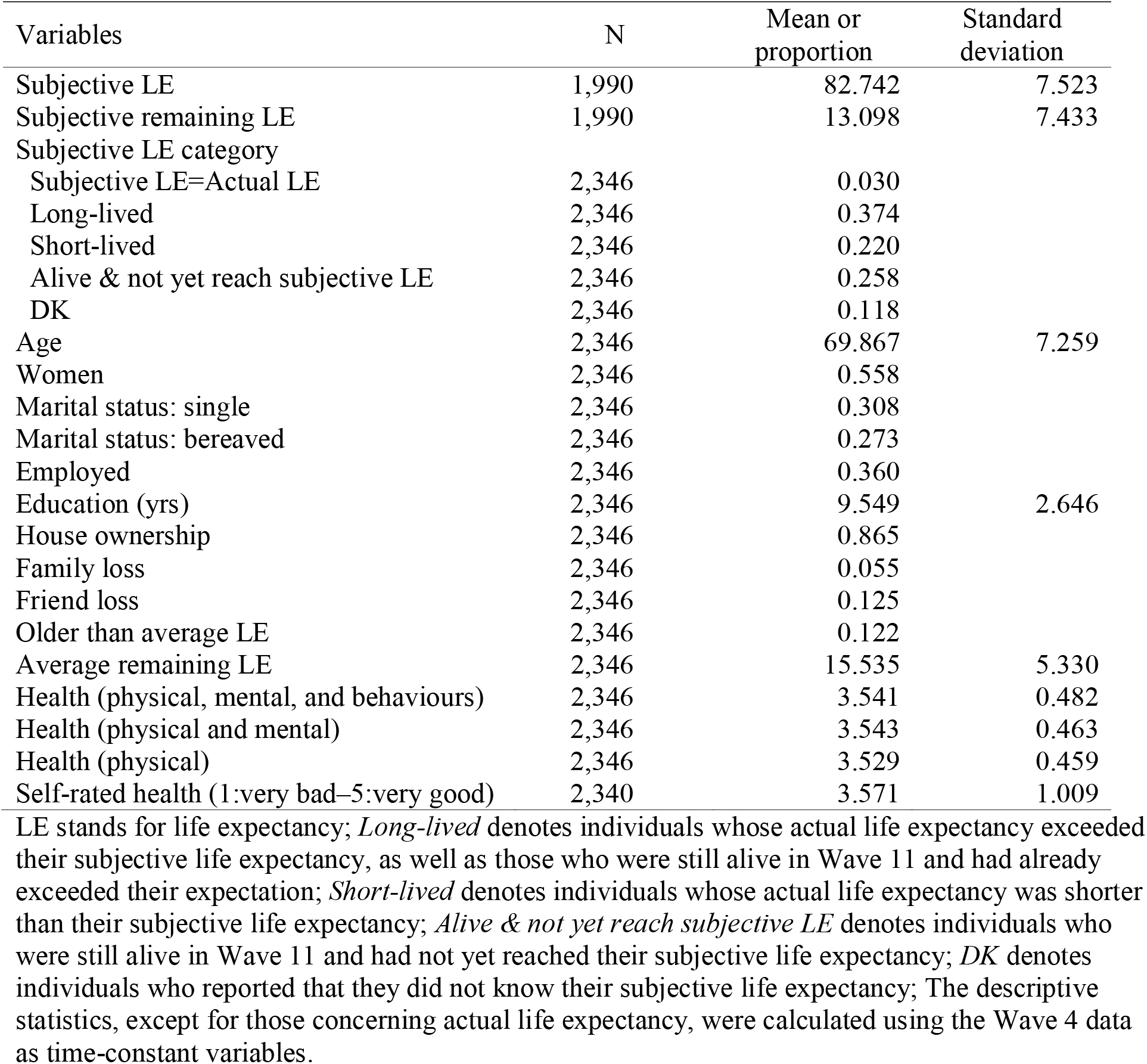
Descriptive statistics of Wave 4 respondents.

Figure 1 illustrates the distributions and differences between SLE and ALE by gender. While the distributions mostly clustered around where SLE and ALE matched, men generally overestimated their lifespan, whereas women tended to underestimate it. Many people reported an SLE of 80 years for both men and women (Appendix Figure A-3), a figure close to the 1996 sex-aggregated average life expectancy at birth. The prediction errors for remaining life expectancy varied considerably, with women particularly underestimating their lifespan compared to official life tables (Appendix Figure A-4).

**Figure 1.**
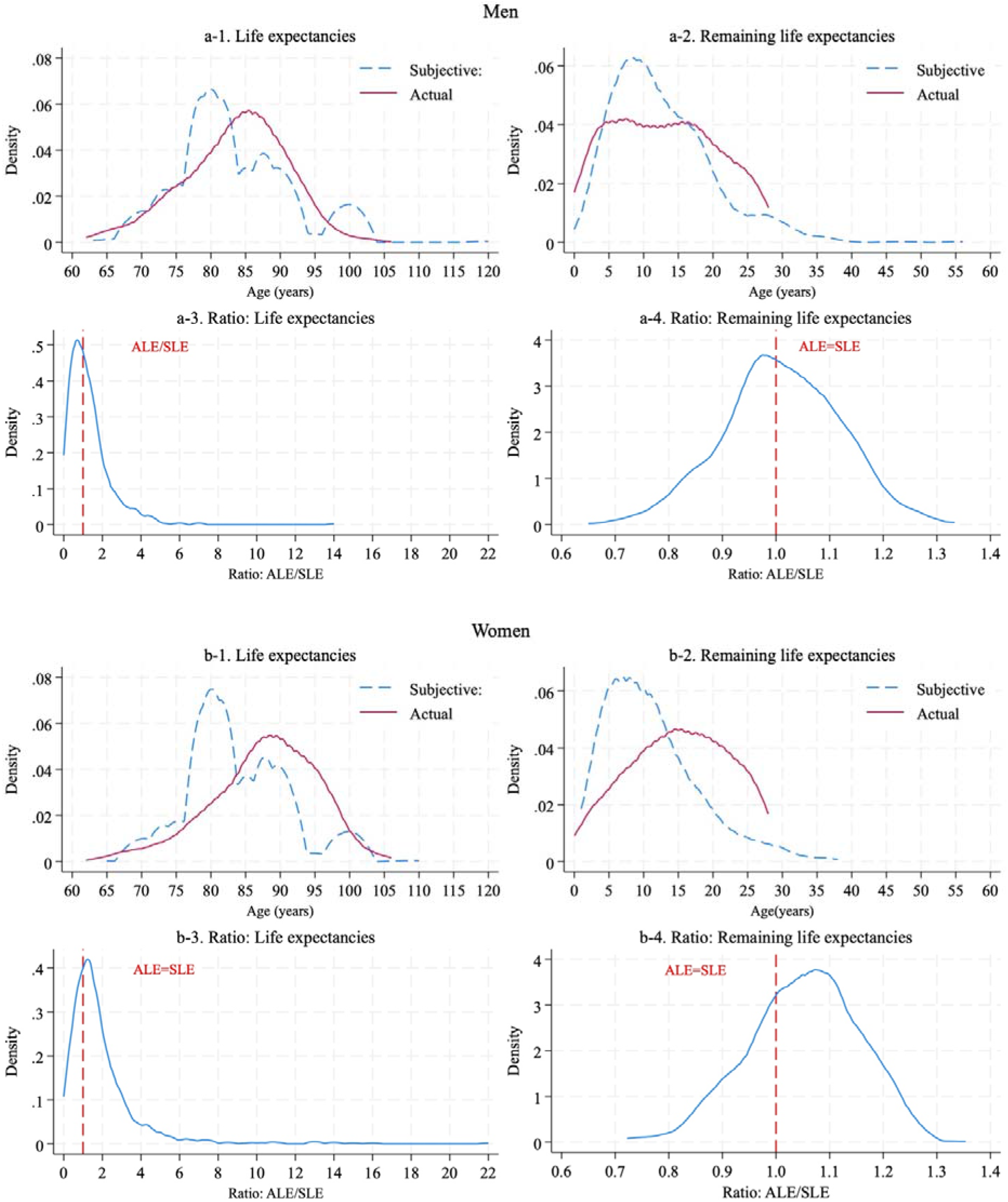
Distributions and discrepancies of subjective and actual life expectancies Note: This visualisation is obtained for the respondents in Wave 4, with both the information on actual and subjective life expectancies available (men: n = 745, women: n = 769). ALE stands for actual life expectancy, while SLE stands for subjective life expectancy.

### 3.2. Predictability of SLE for actual mortality

Figure 2 displays the results of the survival analysis, indicating that a longer SLE correlates with increased survival over a follow-up period of up to 28 years, even after adjusting for other variables. Each additional year in SLE was linked to a hazard ratio of 0.984. Compared to the first quartile, the hazard ratios for the second and third quartiles were 0.836 (cluster-robust SE: 0.059) and 0.804 (0.063) in the basic model adjusted for age and gender, and 0.848 (0.061) and 0.795 (0.063) in the fully adjusted model. Detailed findings are available in Appendix Table A-5.

**Figure 2.**
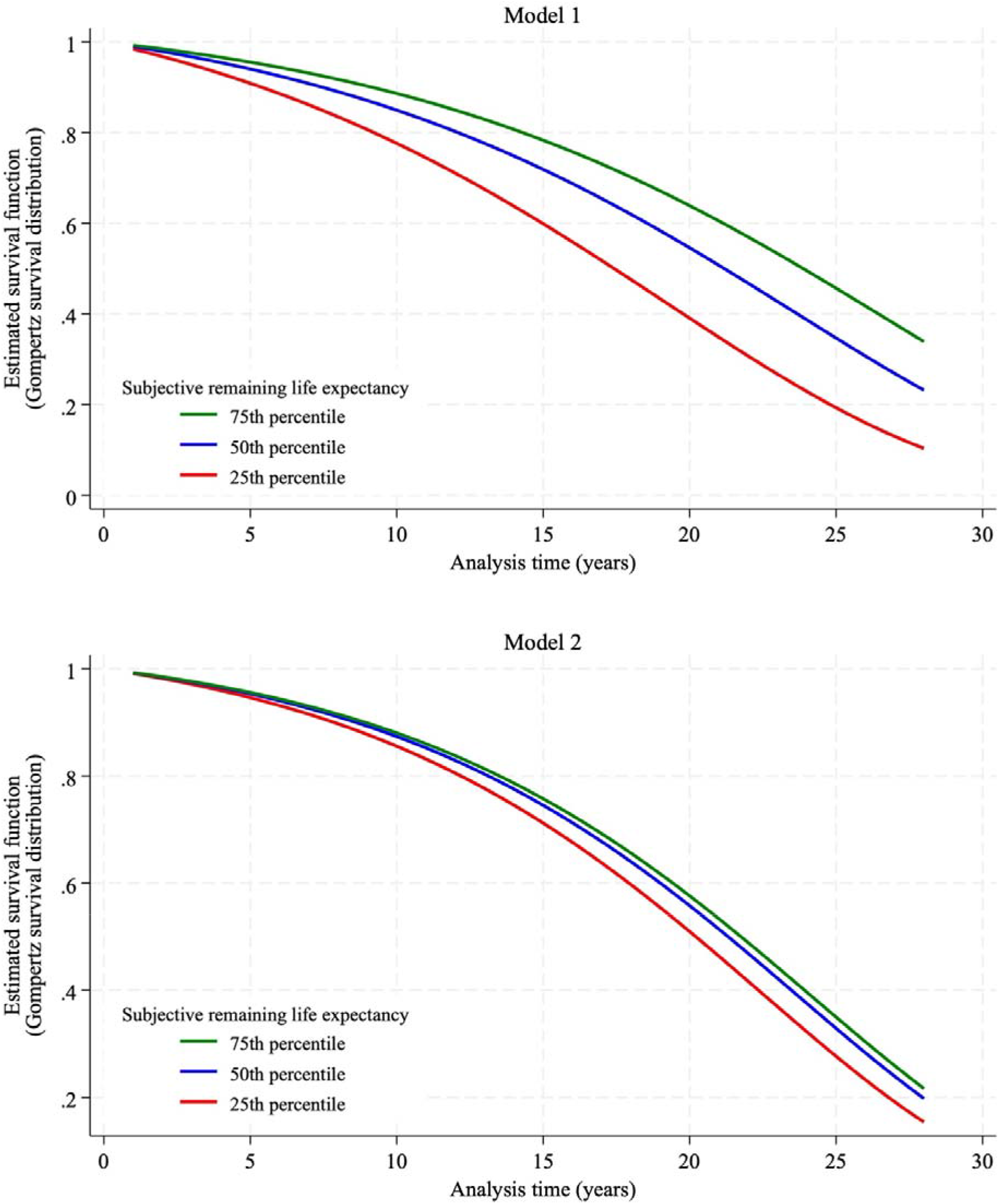
Subjective remaining life expectancy and actual morality between Wave 4 and Wave 11 This visualisation is based on the estimation results in Appendix Table A-5. Model 1 includes controls for age, age², and gender, while Model 2 includes additional controls for education, marital status, employment status, house ownership, experiences of family or friend loss, a dummy variable indicating whether the respondent is older than the sex-specific average life expectancy at birth and the age- and sex-specific average remaining life expectancy. These variables were obtained from the information in Wave 4.

Predicted marginal survival times from the survival analysis indicated that SLE was positively associated with ALE; however, the marginal effects were substantially smaller than one (= 0.119), suggesting that the association between SLE and ALE was inelastic.

### 3.3. Socioeconomic and health disparities

Disparities in both ALE and SLE by education and health were evident (Figure 4), showing a pro-higher-education and pro-health distribution, with CI values from 0.045 to 0.098. The concentration curves reveal that disparities in SLE by education were less marked than those in ALE, while health-related differences were even less noticeable. For accuracy, measured by the ratio of actual to SLE, no educational disparity was observed. However, a health-related disparity was evident, with the concentration index indicating greater concentration among less healthy individuals (CI: −0.072). This implies that individuals in poorer health tend to expect shorter life expectancy.

**Figure 3.**
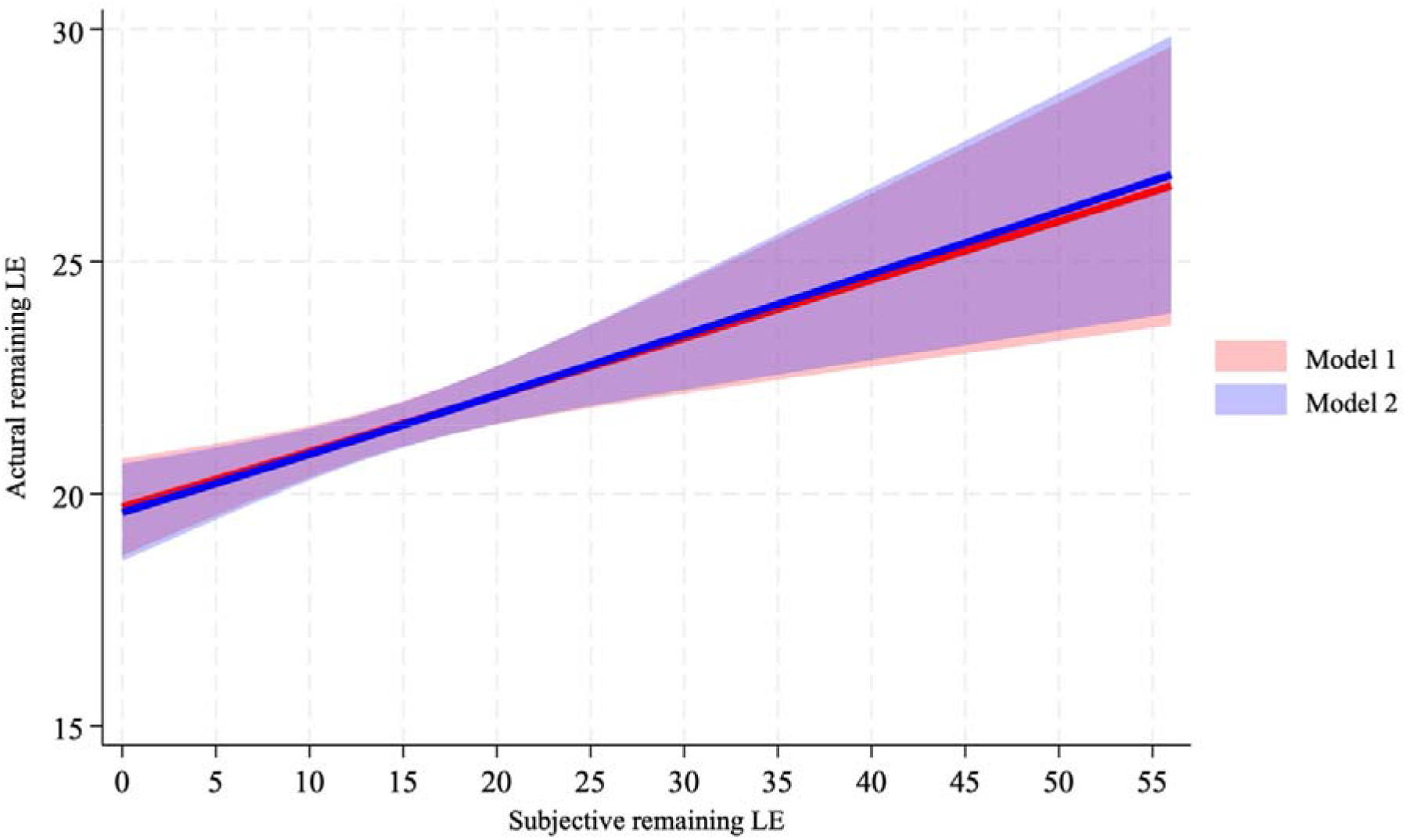
Predicted marginal survival time in relation to subjective remaining life expectancy Note: LE stands for life expectancy. This visualisation is based on the estimation result in Appendix Table A-5. Model 1 refers to the simpler model that includes only age and gender controls, corresponding to Model (1)-1 in the table. Model 2 refers to the fully adjusted model, corresponding to Model (2)-1. The line indicates point estimates, with the shaded area representing the 95% confidence interval.

**Figure 4.**
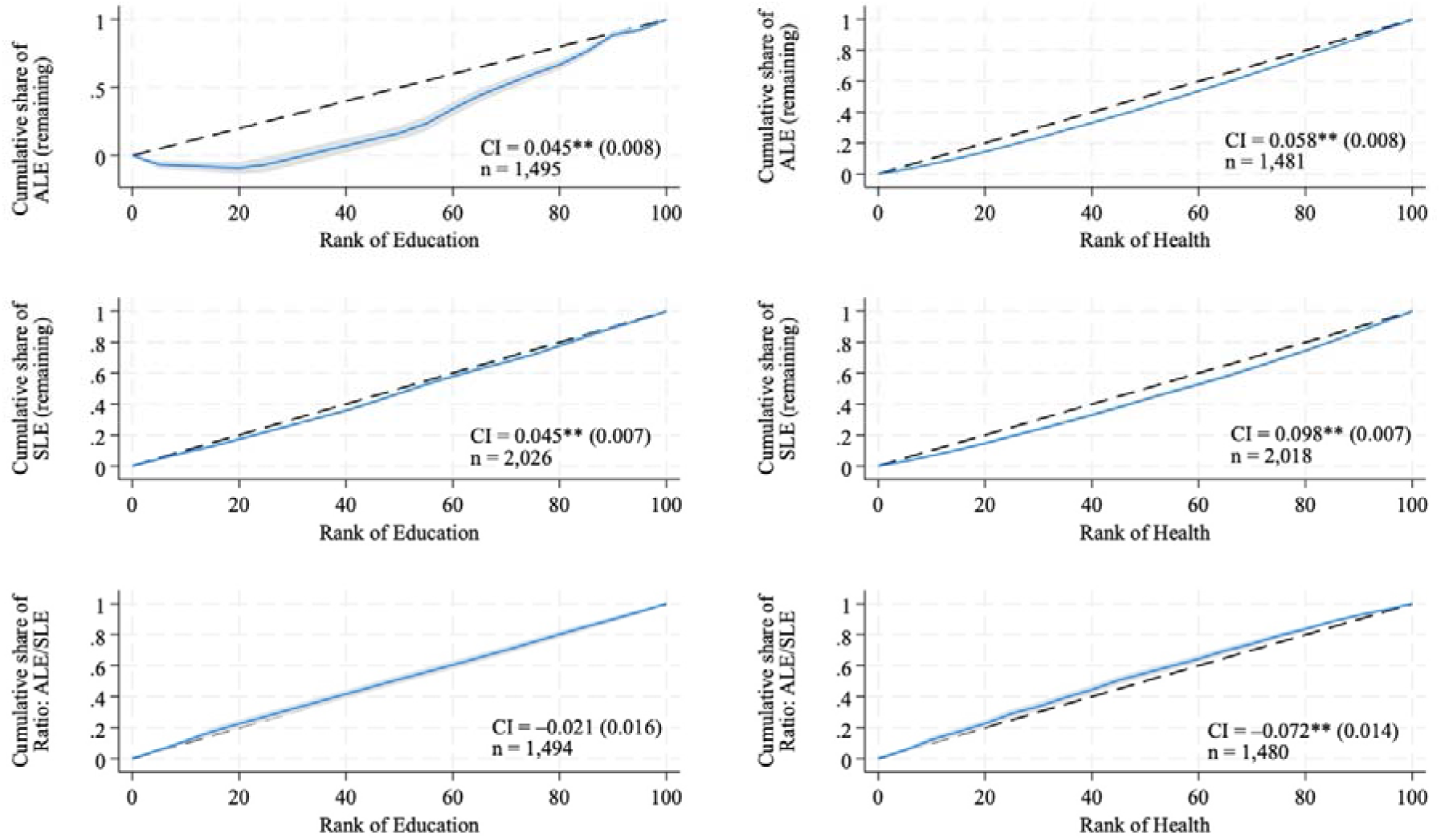
Concentration curves: Disparities in actual and subjective remaining life expectancies by education and health Note: ALE represents actual life expectancy and SLE stands for subjective life expectancy. The ratio (=ALE/SLE) is a continuous variable. Blue lines represent Lorenz curves with shaded areas representing 95% confidence intervals. Health denotes the health score estimated with the information on physical and mental health and health behaviours.

### 3.4. Determinants

Longer SLE was linked to being male, having longer reference ages based on the average remaining life expectancy at current age and gender, and better health status (Appendix Table A-6). In terms of accuracy, women and those with higher education generally lived longer than anticipated, with marginal effects of 0.528 (SE: 0.125) and 0.015 (0.005), respectively (Table 2). The main findings were robust to multiple imputation, but it turned out that individuals with higher education were significantly less likely to report “do not know” to the SLE question (Appendix Table A-7).

**Table 2.** Multinomial logistic regression results: Determinants of the accuracy of survival expectations and “do not know” responses.

|  | Equal | Long-lived | Short-lived | DK |
| --- | --- | --- | --- | --- |
| Age | 0.003<br>(0.020) | -0.046<br>(0.056) | 0.100*<br>(0.050) | -0.057<br>(0.045) |
| Age<sup>2</sup> | 0.002<br>(0.011) | -0.020<br>(0.030) | -0.028<br>(0.027) | 0.046<br>(0.024) |
| Women | -0.019<br>(0.034) | 0.528**<br>(0.125) | -0.494**<br>(0.102) | -0.015<br>(0.088) |
| Health | -0.006<br>(0.012) | 0.008<br>(0.026) | 0.008<br>(0.024) | -0.010<br>(0.021) |
| Education (years) | -0.001<br>(0.002) | 0.015**<br>(0.005) | -0.006<br>(0.005) | -0.008<br>(0.004) |
| Single | 0.017<br>(0.021) | -0.026<br>(0.066) | 0.043<br>(0.060) | -0.034<br>(0.057) |
| Bereaved | -0.032<br>(0.022) | 0.021<br>(0.069) | -0.017<br>(0.062) | 0.028<br>(0.059) |
| Employed | -0.004<br>(0.012) | 0.076**<br>(0.029) | -0.035<br>(0.026) | -0.037<br>(0.024) |
| House ownership | -0.007<br>(0.014) | 0.002<br>(0.037) | 0.010<br>(0.033) | -0.005<br>(0.029) |
| Family death | -0.506**<br>(0.064) | 0.314**<br>(0.061) | 0.098<br>(0.054) | 0.094*<br>(0.046) |
| Friend death | 0.003<br>(0.014) | 0.053<br>(0.037) | -0.007<br>(0.031) | -0.049<br>(0.032) |
| Older than average LE | 0.000<br>(0.023) | 0.038<br>(0.066) | -0.039<br>(0.055) | 0.001<br>(0.050) |
| Average remaining LE | 0.007<br>(0.009) | -0.108**<br>(0.031) | 0.083**<br>(0.026) | 0.018<br>(0.022) |
| Observations |  |  | 1,820 |  |

Note: *Equal* denotes individuals whose actual life expectancy corresponded to their subjective life expectancy; *Long-lived* denotes individuals whose actual life expectancy exceeded their subjective life expectancy, as well as those who were still alive in Wave 11 and had already exceeded their expectation; *Short-lived* denotes individuals whose actual life expectancy was shorter than their subjective life expectancy; *DK* denotes individuals who reported that they did not know their subjective life expectancy; Values are marginal effects with cluster robust standard errors in parentheses; Estimated with the independent variables from Wave 4 (1996) with cross-sectional and longitudinal weights; \*\* p<0.01, \* p<0.05.

Regarding focal point responses at five-year intervals, which may cause divergence between SLE and ALE, these responses were less common among those with higher education levels (Table 3). This trend was also seen in other measures related to body weight and height (Appendix Table A-8). A similar pattern was not observed for responses at age 80; however, the likelihood of reporting this age increased as individuals approached it.

**Table 3.**
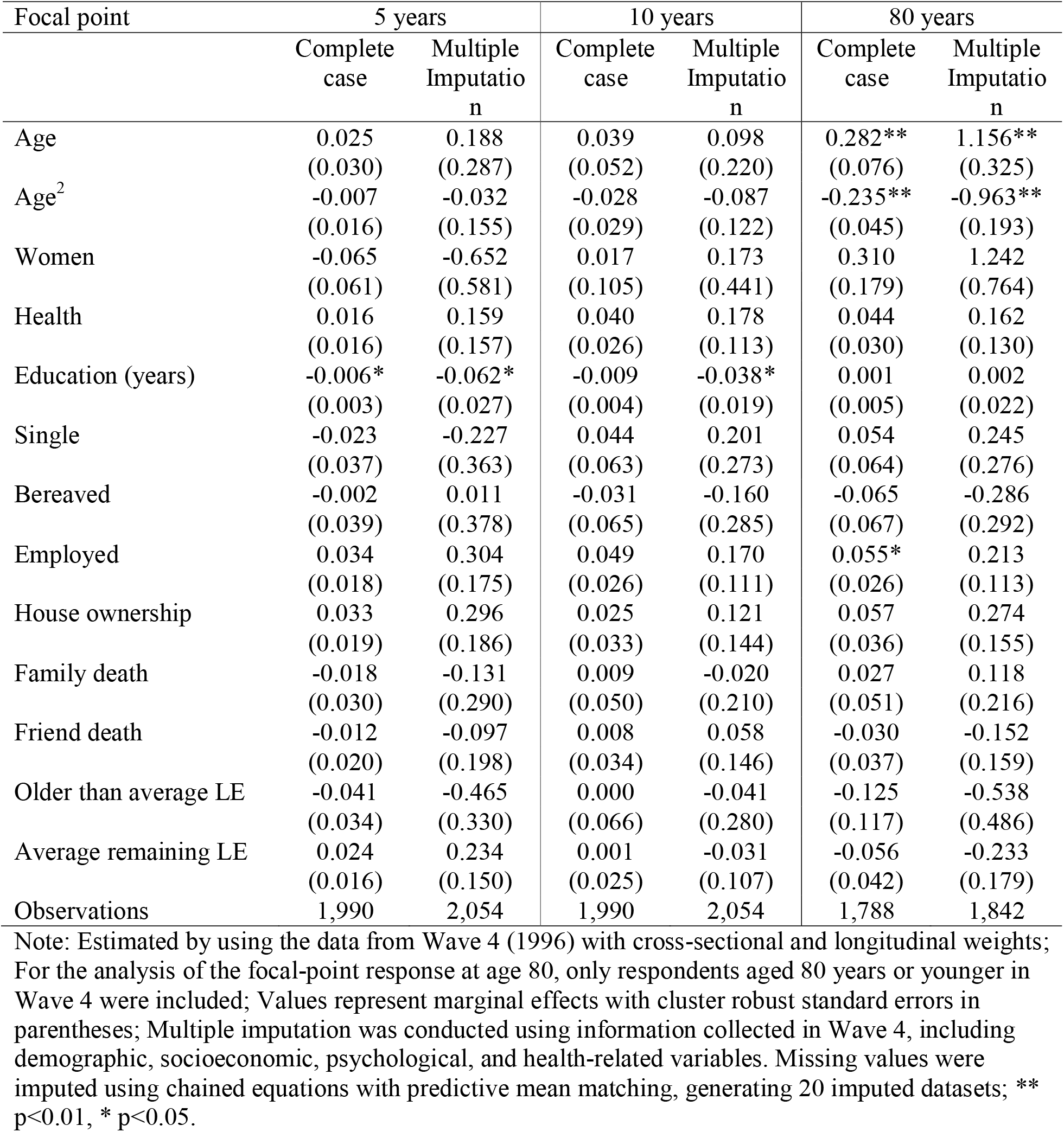
Determinants of focal point responses on subjective life expectancy

## 4. **Discussion and conclusion**

### 4.1 Interpretation

This study analysed SLE using the point-estimate method, evaluating its prognostic value for ALE and determinants among a nationally representative sample of older Japanese with up to 28 years of follow-up. Results show that SLE can predict actual mortality and ALE, but inaccuracies exist, notably a higher number of individuals living beyond their expectations. Women and those with higher education levels generally outlive their SLE, while those in poorer health tended not to do so. Nevertheless, individuals with higher education are less likely to give focal-point responses to the SLE question.

Initially, those with shorter SLE had reduced survival rates, even after accounting for demographic and socioeconomic variables, suggesting that each additional year of SLE corresponded to a 1.6–1.7 percentage-point lower mortality hazard. This aligns with the known link between SLE and mortality. Though the association was inelastic, SLE was still positively related to actual age at death. Supporting prior research (Hurd & McGarry, 2002; van Solinge & Henkens, 2018), our results validate the predictive power of SLE based on the point-estimate method. Since this approach is less burdensome to report than the subjective probability method, our findings offer valuable insights into capturing individuals’ lifespan expectations.

Second, despite significant associations between SLE and actual mortality and ALE, substantial and heterogeneous divergence remains. Most respondents lived longer than they expected, with responses mainly clustered around age 80, close to the sex-aggregated average life expectancy at birth. This is similar to the previous findings that older Americans tend to underestimate their survival probabilities and have inaccurate subjective expectations (Bago d’Uva et al., 2020; Elder, 2013). One explanation is that younger individuals rely on the average life expectancy at birth when estimating SLE, then adjust this estimate upwards over time based on other information—such as family members’ ages at death or milestones (e.g. 100 years), which might reflect aspirations rather than realistic expectations. When referencing an average value, it is better to use sex- and age-specific remaining life expectancy instead of the overall life expectancy at birth. However, many individuals may anchor their SLE to this salient measure, leading to potentially biased, shorter expectations.

The same reasoning also applies to the finding that women and individuals with higher education tend to outlive their SLE. Women generally live longer than men (Patwardhan et al., 2024), and higher education correlates with longer life expectancy (Balaj et al., 2024). Broad averages may obscure longevity benefits linked to these groups’ characteristics. Another explanation for the gender differences could be that women might have lower optimism than men in assessing their mortality risks (Karmarkar, 2023), which might cause them to underestimate their actual lifespans.

Considering the link between education and focal-point and do-not-know responses, this might have influenced the variation in SLE’s predictive validity due to bounded rationality. People with less education and lower cognitive ability tend to be less accurate in estimating their survival chances (Bago d’Uva et al., 2020). Relatedly, it has been reported that the predictive value of self-rated health for mortality varies by education (Dowd & Zajacova, 2007). Nevertheless, those with higher education levels often gave inaccurate predictions, frequently anticipating shorter lifespans than their true ones.

#### Policy implications

Differences between SLE and ALE can cause suboptimal economic behaviours, affecting decisions about retirement, social security benefits, saving, and wealth decumulation. Underestimating life expectancy may lead to insufficient retirement funds, while overestimating it could result in unnecessary frugality or even poverty. Addressing bounded rationality—such as through better financial literacy and consumer-friendly savings plans—and bounded willpower—by applying behavioural insights to service packages (Thaler & Benartzi, 2004)—is crucial. Additionally, guiding individuals with more accurate information about their SLE is important. This includes helping them estimate life expectancy based on sex- and age-specific averages, considering the link between their age at death and that of their same-sex parent (excluding accidental deaths), and acknowledging survival disparities linked to socioeconomic and health factors.

Japan’s current public old-age pension scheme allows individuals to choose when to claim benefits between the ages of 60–75, with 65 typically serving as the standard eligibility age. Claiming early results in a 0.4% reduction per month, while deferring benefits yields an extra 0.7% monthly increase. These adjustments apply for life and are fixed once made. As of March 2025, 23.2% of eligible individuals claimed benefits before age 65, and 2.4% deferred for the basic pension; meanwhile, only 1.2% claimed early and 1.9% deferred for the earnings-related pension (Ministry of Health, Labor and Welfare, 2026). Early claimers of the basic pension tend to have higher mortality rates, which might justify their behaviour (Ministry of Health, Labor and Welfare, 2019). However, if these actions are based on misinformed expectations, corrective measures are necessary. Providing information on age- and sex-specific life expectancy and the potential gains or losses from early claiming or deferring could help individuals make more informed decisions.

#### Limitations

This study has several limitations. First, SLE was only measured once in Wave 4, so we were unable to account for changes over time. As people gain new information, such as diagnosis of illnesses, medical and drug advancements, or the death of loved ones, their lifespan expectations might shift. In our sample of older adults, many may have already experienced more events influencing their SLE than younger individuals. However, their SLE can still vary, especially as they near death. Second, we analysed a sample of older adults only. Future research should explore whether people can accurately predict their SLE, particularly in early and middle adulthood when long-term financial planning and retirement preparations are crucial. Third, the survey lacked detailed data on parental longevity (e.g. the age at death of the same-sex parent), which is positively associated with SLE (van Solinge & Henkens, 2018). While the survey did note experiences related to the death of family or close friends, not including this critical information might have resulted in omitted variable bias.

In summary, SLE is a useful indicator for predicting actual mortality and ALE. Nonetheless, differences between SLE and ALE are significant. Accuracy differs among individuals and is linked to gender and socioeconomic factors, with women and those with higher education generally living longer than expected. Focal-point responses to the SLE question, which are more frequent among less educated individuals, also seem to play a role in this inaccuracy. Providing personalised information, such as age- and sex-specific average remaining life expectancy instead of overall average at birth, could help individuals develop more precise expectations and improve their economic decisions.

## Supplementary materials for

Appendix Table A-1: Descriptive statistics for health score estimation

|  | Variables | Mean or proportion | Standard deviation |
| --- | --- | --- | --- |
| Dependent variable | Self-rated health | 3.430 | 1.009 |
| Demographic factors | Age | 74.375 | 7.568 |
|  | Women | 0.553 | 0.497 |
| Physical health | Body mass index | 22.482 | 3.321 |
|  | Chronic respiratory disease | 0.063 | 0.243 |
|  | Chronic respiratory disease (DK) | 0.012 | 0.110 |
|  | Hypertension | 0.362 | 0.481 |
|  | Hypertension (DK) | 0.008 | 0.090 |
|  | Heart disease | 0.137 | 0.344 |
|  | Heart disease (DK) | 0.011 | 0.105 |
|  | Diabetes | 0.103 | 0.304 |
|  | Diabetes (DK) | 0.012 | 0.107 |
|  | Cerebrovascular disease | 0.045 | 0.208 |
|  | Cerebrovascular disease (DK) | 0.014 | 0.118 |
|  | Cancer | 0.039 | 0.193 |
|  | Cancer (DK) | 0.015 | 0.121 |
|  | Chronic lower back pain | 0.215 | 0.411 |
|  | Chronic lower back pain (DK) | 0.010 | 0.099 |
|  | Arthritis | 0.154 | 0.361 |
|  | Arthritis (DK) | 0.012 | 0.108 |
|  | ADL disability | 0.368 | 1.645 |
|  | IADL disability | 0.660 | 2.248 |
|  | Difficulty: continue to stand up | 0.121 | 0.326 |
|  | Difficulty: continue to stand up (DK) | 0.004 | 0.062 |
|  | Difficulty: walk 200-300m | 0.122 | 0.327 |
|  | Difficulty: walk 200-300m (DK) | 0.003 | 0.051 |
|  | Difficulty: squat down | 0.173 | 0.378 |
|  | Difficulty: squat down (DK) | 0.004 | 0.064 |
|  | Difficulty: continue to stand up | 0.124 | 0.330 |
|  | Difficulty: continue to stand up (DK) | 0.005 | 0.068 |
|  | Difficulty: use fingers | 0.042 | 0.201 |
|  | Difficulty: use fingers (DK) | 0.003 | 0.053 |
|  | Difficulty: carry heavy things | 0.243 | 0.429 |
|  | Difficulty: carry heavy things (DK) | 0.010 | 0.102 |
|  | Difficulty: climb two or three steps | 0.117 | 0.322 |
|  | Difficulty: climb two or three steps (DK) | 0.004 | 0.064 |
| Mental health | CES-D | 3.181 | 3.265 |
|  | Cognitive functioning | 1.096 | 1.302 |
| Health behaviours | Smoking | 0.133 | 0.340 |
|  | Smoking (DK) | 0.000 | 0.017 |
|  | Alcohol consumption | 0.384 | 0.486 |
|  | Alcohol consumption (DK) | 0.000 | 0.019 |
|  | Habit: gardening | 0.700 | 0.458 |
|  | Habit: gardening (DK) | 0.004 | 0.066 |
|  | Habit: exercise | 0.561 | 0.496 |
|  | Habit: exercise (DK) | 0.003 | 0.051 |
|  | Habit: walk | 0.639 | 0.480 |
|  | Habit: walk (DK) | 0.002 | 0.049 |

DK: Do not know / No answer; These descriptive statistics summarise responses from 20,400 observations by 6,823 unique individuals between Wave 1 and Wave 11.

Appendix Table A-2: Health score estimation by the random-effects ordered probit model

| Dependent variable: Self-rated health | Physical, mental, behaviours | Physical + mental | Physical only |
| --- | --- | --- | --- |
| Age | -0.386** (0.040) | -0.380** (0.040) | -0.390** (0.040) |
| Age<sup>2</sup> | 0.267** (0.027) | 0.262** (0.027) | 0.263** (0.027) |
| Women | 0.277** (0.049) | 0.095* (0.045) | 0.039 (0.045) |
| Body mass index | 0.040** (0.006) | 0.039** (0.006) | 0.039** (0.006) |
| Chronic respiratory disease | -0.764** (0.077) | -0.792** (0.078) | -0.833** (0.079) |
| Chronic respiratory disease (DK) | -0.089 (0.263) | -0.070 (0.272) | -0.076 (0.275) |
| Hypertension | -0.498** (0.039) | -0.487** (0.039) | -0.494** (0.039) |
| Hypertension (DK) | -0.060 (0.215) | -0.130 (0.215) | -0.205 (0.218) |
| Heart disease | -0.844** (0.054) | -0.880** (0.054) | -0.894** (0.054) |
| Heart disease (DK) | -0.126 (0.208) | -0.067 (0.207) | -0.026 (0.212) |
| Diabetes | -0.678** (0.064) | -0.680** (0.065) | -0.695** (0.065) |
| Diabetes (DK) | -0.383 (0.220) | -0.351 (0.222) | -0.363 (0.222) |
| Cerebrovascular disease | -0.620** (0.093) | -0.637** (0.094) | -0.702** (0.094) |
| Cerebrovascular disease (DK) | -0.109 (0.200) | -0.069 (0.197) | -0.119 (0.198) |
| Cancer | -0.712** (0.105) | -0.718** (0.106) | -0.766** (0.108) |
| Cancer (DK) | -0.006 (0.205) | -0.065 (0.209) | -0.106 (0.209) |
| Chronic lower back pain | -0.479** (0.044) | -0.469** (0.044) | -0.504** (0.044) |
| Chronic lower back pain (DK) | -0.012 (0.239) | -0.006 (0.239) | 0.084 (0.239) |
| Arthritis | -0.484** (0.048) | -0.465** (0.048) | -0.477** (0.048) |
| Arthritis (DK) | 0.111 (0.213) | 0.084 (0.211) | 0.133 (0.212) |
| ADL disability | -0.078** (0.020) | -0.083** (0.020) | -0.092** (0.020) |
| ADL disability | -0.056** (0.015) | -0.073** (0.015) | -0.078** (0.015) |
| Difficulty: continue to stand up | -0.341** (0.076) | -0.388** (0.076) | -0.380** (0.076) |
| Difficulty: continue to stand up (DK) | 0.253 (0.285) | 0.237 (0.284) | 0.286 (0.289) |
| Difficulty: walk 200-300m | -0.317** (0.079) | -0.391** (0.079) | -0.400** (0.080) |
| Difficulty: walk 200-300m (DK) | 0.304 (0.378) | 0.267 (0.377) | 0.347 (0.370) |
| Difficulty: squat down | -0.374** (0.059) | -0.395** (0.059) | -0.419** (0.058) |
| Difficulty: squat down (DK) | -0.274 (0.300) | -0.255 (0.295) | -0.252 (0.292) |
| Difficulty: continue to stand up | -0.298** (0.068) | -0.336** (0.069) | -0.352** (0.069) |
| Difficulty: continue to stand up (DK) | 0.053 (0.254) | 0.074 (0.262) | 0.015 (0.262) |
| Difficulty: use fingers | -0.356** (0.094) | -0.357** (0.095) | -0.411** (0.095) |
| Difficulty: use fingers (DK) | 0.179 (0.409) | 0.124 (0.421) | 0.193 (0.415) |
| Difficulty: carry heavy things | -0.709** (0.052) | -0.779** (0.052) | -0.800** (0.051) |
| Difficulty: carry heavy things (DK) | -0.615** (0.173) | -0.689** (0.172) | -0.668** (0.174) |
| Difficulty: climb two or three steps | -0.228** (0.070) | -0.255** (0.071) | -0.263** (0.071) |
| Difficulty: climb two or three steps (DK) | 0.103 (0.265) | 0.026 (0.266) | 0.039 (0.276) |
| CES-D | -0.080** (0.006) | -0.076** (0.006) |  |
| Cognitive functioning | -0.057** (0.014) | -0.076** (0.014) |  |
| Smoking | 0.215** (0.058) |  |  |
| Smoking (DK) | 0.561 (1.307) |  |  |
| Alcohol consumption | 0.436** (0.043) |  |  |
| Alcohol consumption (DK) | 0.006 (1.323) |  |  |
| Habit: gardening | 0.336** (0.042) |  |  |
| Habit: gardening (DK) | -0.476 (0.268) |  |  |
| Habit: exercise | 0.318** (0.037) |  |  |
| Habit: exercise (DK) | 0.451 (0.444) |  |  |
| Habit: walk | 0.197** (0.039) |  |  |
| Habit: walk (DK) | 1.257* (0.536) |  |  |
| Observations (Number of individuals) | 20,400 (6,823) | 20,416 (6,825) | 20,489 (6,834) |

Note: Values are coefficients with cluster robust standard errors in parentheses; \*\* p<0.01, \* p<0.05; Estimated using the data from Wave 1 (1987) to Wave 11 (2024) with cross-sectional and longitudinal weights.

Appendix Table A-3: Valid responses and response rates

|  | Newly added | Continuous | Total |
| --- | --- | --- | --- |
| Wave 1 (1987) | 2,200 (67.2%) | N.A. | 2,200 (67.2%) |
| Wave 2 (1990) | 366 (69.9%) | 1,671 (89.5%) | 2,037 (73.3%) |
| Wave 3 (1993) | N.A. | 1,864 (92.6%) | 1,864 (92.6%) |
| Wave 4 (1996) | 898 (80.9%) | 1,549 (89.1%) | 2,447 (86.0%) |
| Wave 5 (1999) | 1,635 (82.6%) | 2,354 (87.2%) | 3,989 (85.2%) |
| Wave 6 (2002) | 1,246 (85.9%) | 1,999 (82.4%) | 3,245 (83.7%) |
| Wave 7 (2006) | N.A. | 2,459 (75.4%) | 2,459 (75.4%) |
| Wave 8 (2012) | 1,450 (58.9%) | 1,490 (78.3%) | 2,940 (67.4%) |
| Wave 9 (2017) | N.A. | 1,748 (77.4%) | 1,748 (77.4%) |
| Wave 10 (2021) | 1,227 (45.9%) | 706 (77.1%) | 1,933 (53.9%) |
| Wave 11 (2024) | N.A. | 1,329 (72.7%) | 1,329 (72.7%) |

Note: Individuals whose deaths or admissions to care institutions had been confirmed prior to the surveys were excluded.

Appendix Table A-4. Comparison of alive and deceased samples from Wave 4 to Wave 11

|  | Alive |  |  | Deceased |  |  |
| --- | --- | --- | --- | --- | --- | --- |
| Age | 606 | 64.726 | 4.228 | 1,740 | 71.657 | 7.240 |
| Women | 606 | 0.675 | 0.469 | 1,740 | 0.517 | 0.500 |
| Education (years) | 606 | 10.398 | 2.628 | 1,740 | 9.253 | 2.589 |
| House ownership | 606 | 0.870 | 0.337 | 1,740 | 0.864 | 0.343 |
| Marital status: single | 606 | 0.215 | 0.411 | 1,740 | 0.341 | 0.474 |
| Marital status: bereaved | 606 | 0.180 | 0.384 | 1,740 | 0.305 | 0.461 |
| Employed | 606 | 0.470 | 0.500 | 1,740 | 0.322 | 0.467 |
| Family loss | 606 | 0.059 | 0.237 | 1,740 | 0.053 | 0.225 |
| Friend loss | 606 | 0.091 | 0.288 | 1,740 | 0.137 | 0.344 |
| Health score (physical, mental, and behaviours) | 606 | 3.734 | 0.336 | 1,740 | 3.473 | 0.507 |
| Health score (physical and mental) | 606 | 3.736 | 0.307 | 1,740 | 3.476 | 0.488 |
| Health score (physical) | 606 | 3.727 | 0.304 | 1,740 | 3.460 | 0.483 |
| Self-rated health | 602 | 3.766 | 0.962 | 1,738 | 3.503 | 1.016 |
| Subjective remaining LE | 526 | 16.279 | 7.368 | 1,464 | 11.956 | 7.119 |
| Subjective LE | 526 | 80.791 | 6.881 | 1,464 | 83.443 | 7.621 |

Note: LE stands for life expectancy.

Appendix Table A-5. Prognostic value of subjective remaining life expectancy for actual mortality: survival analysis results

|  | (1) - 1 | (1) - 2 | (2) - 1 | (2) - 2 |
| --- | --- | --- | --- | --- |
| Subjective remaining LE (cont.) | 0.984**<br>(0.005) |  | 0.983**<br>(0.005) |  |
| Subjective remaining LE (Q1) |  | Ref. |  | Ref. |
| Subjective remaining LE (Q2) |  | 0.836*<br>(0.059) |  | 0.848*<br>(0.061) |
| Subjective remaining LE (Q3) |  | 0.804**<br>(0.063) |  | 0.795**<br>(0.063) |
| Age | 1.471**<br>(0.112) | 1.470**<br>(0.114) | 1.334*<br>(0.168) | 1.334*<br>(0.168) |
| Age<sup>2</sup> | 0.836**<br>(0.044) | 0.837**<br>(0.044) | 0.865*<br>(0.061) | 0.865*<br>(0.061) |
| Women | 0.480**<br>(0.027) | 0.483**<br>(0.028) | 0.524**<br>(0.122) | 0.535**<br>(0.125) |
| Education (years) |  |  | 0.932**<br>(0.011) | 0.934**<br>(0.011) |
| House ownership |  |  | 1.007<br>(0.088) | 1.007<br>(0.088) |
| Marital status: single |  |  | 1.392*<br>(0.216) | 1.381*<br>(0.213) |
| Marital status: bereaved |  |  | 0.698*<br>(0.113) | 0.700*<br>(0.113) |
| Employed |  |  | 0.845*<br>(0.056) | 0.854*<br>(0.057) |
| Family death |  |  | 0.985<br>(0.121) | 0.973<br>(0.121) |
| Friend death |  |  | 1.048<br>(0.084) | 1.046<br>(0.084) |
| Older than average LE at birth |  |  | 1.239<br>(0.159) | 1.252<br>(0.161) |
| Average remaining LE |  |  | 0.955<br>(0.057) | 0.953<br>(0.057) |
| gamma | 1.127**<br>(0.005) | 1.126**<br>(0.005) | 1.129**<br>(0.005) | 1.129**<br>(0.005) |
| Constant | 0.000**<br>(0.000) | 0.000**<br>(0.000) | 0.000*<br>(0.000) | 0.000*<br>(0.000) |
| Observations | 2,032 | 2,032 | 2,003 | 2,003 |

Note: Weighted by cross-sectional and longitudinal weights; Values are hazard ratios with cluster robust standard errors in parentheses; Independent variables were based on the information in Wave 4 and treated as time-constant; The Gompertz survival distribution was chosen based on its model fit as assessed by Akaike's Information Criterion; \*\* p<0.01, \* p<0.05; Health-related variables were not included in the models to avoid strong collinearity with subjective life expectancy.

Appendix Table A-6. Determinants of remaining subjective life expectancy

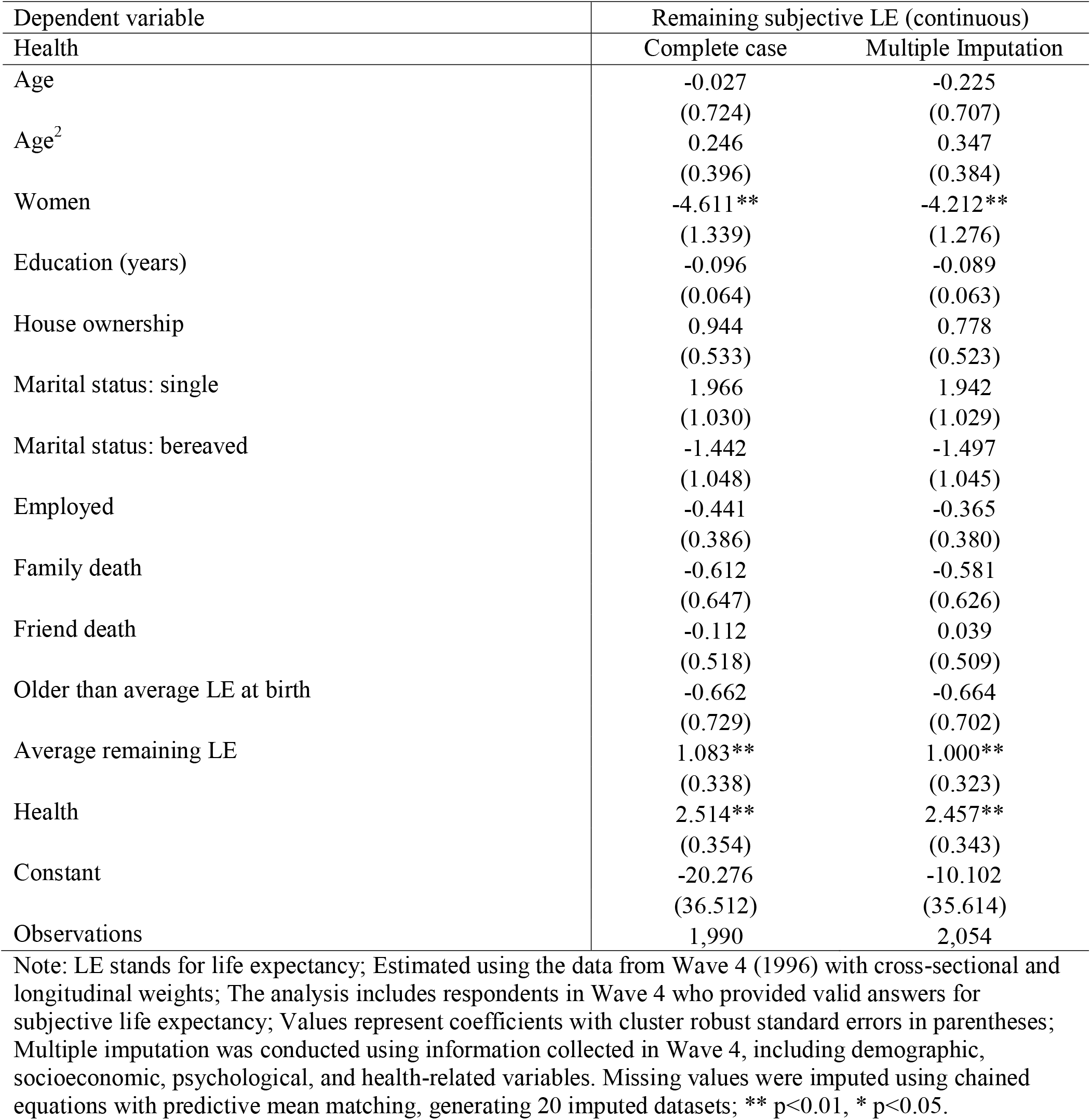

Appendix Table A-7. Multinomial logistic regression results with multiple imputation: Determinants of longer- and shorter-than-expected survival and “do not know” responses

| Ref: Long-lived | Equal | Long-lived | Short-lived | DK |
| --- | --- | --- | --- | --- |
| Age | 0.003<br>(0.020) | -0.001<br>(0.054) | 0.091<br>(0.048) | -0.093*<br>(0.044) |
| Age<sup>2</sup> | 0.002<br>(0.011) | -0.048<br>(0.029) | -0.020<br>(0.026) | 0.066**<br>(0.023) |
| Women | -0.014<br>(0.032) | 0.488**<br>(0.123) | -0.510**<br>(0.097) | 0.037<br>(0.086) |
| Health | -0.007<br>(0.011) | 0.017<br>(0.025) | 0.014<br>(0.023) | -0.024<br>(0.021) |
| Education (years) | -0.001<br>(0.002) | 0.015**<br>(0.005) | -0.006<br>(0.004) | -0.009*<br>(0.004) |
| Single | 0.017<br>(0.021) | -0.026<br>(0.065) | 0.043<br>(0.059) | -0.034<br>(0.056) |
| Bereaved | -0.032<br>(0.022) | 0.023<br>(0.068) | -0.013<br>(0.062) | 0.022<br>(0.058) |
| Employed | -0.004<br>(0.012) | 0.072*<br>(0.029) | -0.035<br>(0.026) | -0.033<br>(0.024) |
| House ownership | -0.005<br>(0.014) | 0.006<br>(0.036) | 0.009<br>(0.032) | -0.010<br>(0.029) |
| Family death | -0.504**<br>(0.063) | 0.312**<br>(0.059) | 0.099<br>(0.053) | 0.092*<br>(0.046) |
| Friend death | 0.002<br>(0.014) | 0.038<br>(0.036) | 0.006<br>(0.030) | -0.045<br>(0.032) |
| Older than average LE | 0.003<br>(0.021) | 0.062<br>(0.064) | -0.063<br>(0.053) | -0.003<br>(0.048) |
| Average remaining LE | 0.006<br>(0.009) | -0.099**<br>(0.031) | 0.087**<br>(0.025) | 0.007<br>(0.022) |
| Observations | 1,907 |  |  |  |

Note: Estimated with the data from Wave 4 (1996) with cross-sectional and longitudinal weights; *Equal* denotes individuals whose actual life expectancy corresponded to their subjective life expectancy; *Long-lived* denotes individuals whose actual life expectancy exceeded their subjective life expectancy, as well as those who were still alive in Wave 11 and had already exceeded their expectation; *Short-lived* denotes individuals whose actual life expectancy was shorter than their subjective life expectancy; *DK* denotes individuals who reported that they did not know their subjective life expectancy; Multiple imputation was conducted using information collected in Wave 4, including demographic, socioeconomic, psychological, and health-related variables. Missing values were imputed using chained equations with predictive mean matching, generating 20 imputed datasets; Values are marginal effects with cluster robust standard errors in parentheses; \*\* p<0.01, \* p<0.05.

Appendix Table A-8. Determinants of focal point responses on body weight and height

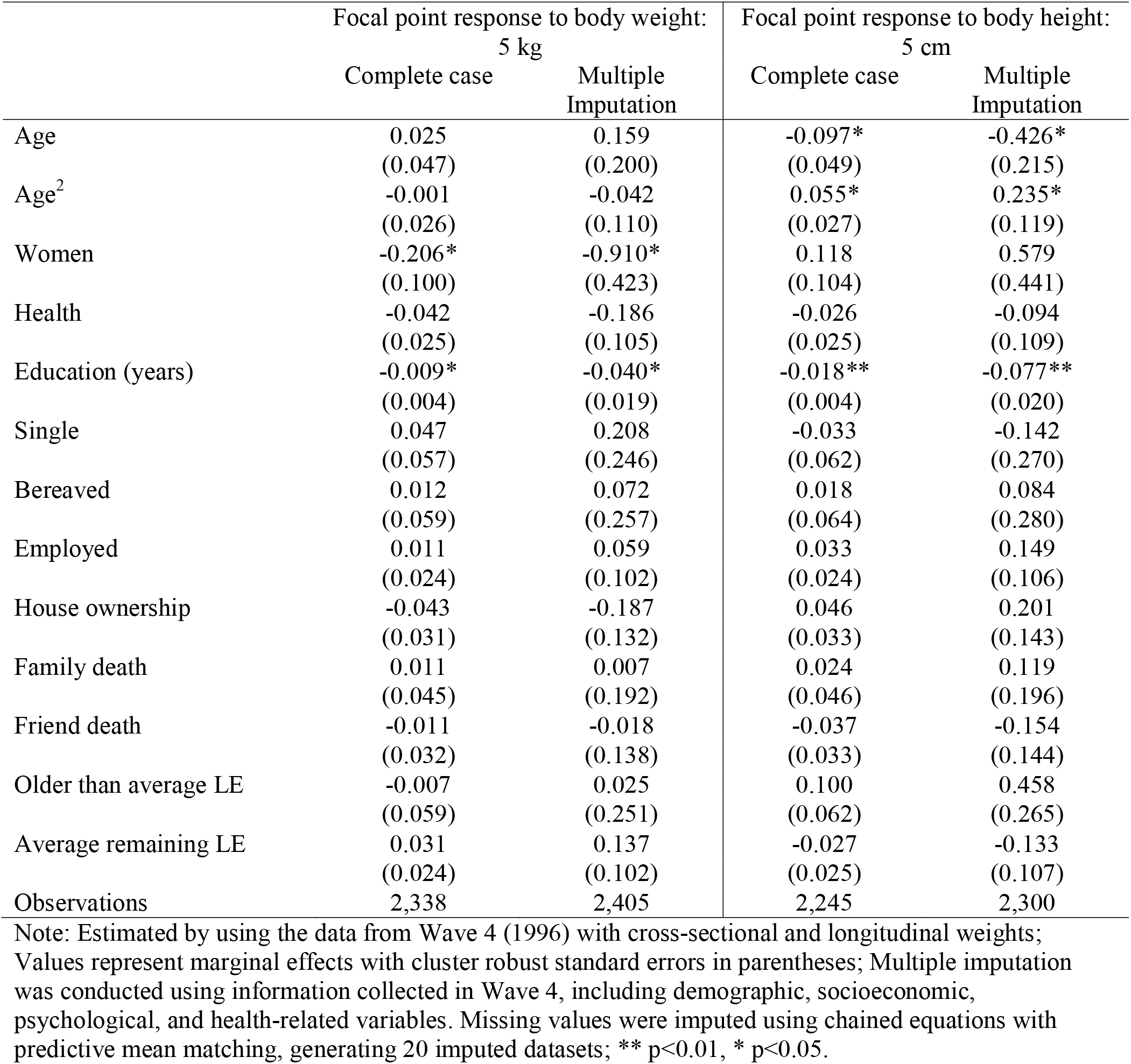

|  | Focal point response to body weight:<br>5 kg |  | Focal point response to body height:<br>5 cm |  |
| --- | --- | --- | --- | --- |
|  | Complete case | Multiple Imputation | Complete case | Multiple Imputation |
| Age | 0.025<br>(0.047) | 0.159<br>(0.200) | -0.097*<br>(0.049) | -0.426*<br>(0.215) |
| Age<sup>2</sup> | -0.001<br>(0.026) | -0.042<br>(0.110) | 0.055*<br>(0.027) | 0.235*<br>(0.119) |
| Women | -0.206*<br>(0.100) | -0.910*<br>(0.423) | 0.118<br>(0.104) | 0.579<br>(0.441) |
| Health | -0.042<br>(0.025) | -0.186<br>(0.105) | -0.026<br>(0.025) | -0.094<br>(0.109) |
| Education (years) | -0.009*<br>(0.004) | -0.040*<br>(0.019) | -0.018**<br>(0.004) | -0.077**<br>(0.020) |
| Single | 0.047<br>(0.057) | 0.208<br>(0.246) | -0.033<br>(0.062) | -0.142<br>(0.270) |
| Bereaved | 0.012<br>(0.059) | 0.072<br>(0.257) | 0.018<br>(0.064) | 0.084<br>(0.280) |
| Employed | 0.011<br>(0.024) | 0.059<br>(0.102) | 0.033<br>(0.024) | 0.149<br>(0.106) |
| House ownership | -0.043<br>(0.031) | -0.187<br>(0.132) | 0.046<br>(0.033) | 0.201<br>(0.143) |
| Family death | 0.011<br>(0.045) | 0.007<br>(0.192) | 0.024<br>(0.046) | 0.119<br>(0.196) |
| Friend death | -0.011<br>(0.032) | -0.018<br>(0.138) | -0.037<br>(0.033) | -0.154<br>(0.144) |
| Older than average LE | -0.007<br>(0.059) | 0.025<br>(0.251) | 0.100<br>(0.062) | 0.458<br>(0.265) |
| Average remaining LE | 0.031<br>(0.024) | 0.137<br>(0.102) | -0.027<br>(0.025) | -0.133<br>(0.107) |
| Observations | 2,338 | 2,405 | 2,245 | 2,300 |

Note: Estimated by using the data from Wave 4 (1996) with cross-sectional and longitudinal weights; Values represent marginal effects with cluster robust standard errors in parentheses; Multiple imputation was conducted using information collected in Wave 4, including demographic, socioeconomic, psychological, and health-related variables. Missing values were imputed using chained equations with predictive mean matching, generating 20 imputed datasets; \*\* p<0.01, \* p<0.05.

Appendix Figure A-1. Kernel density plots for self-rated health and predicted health scores

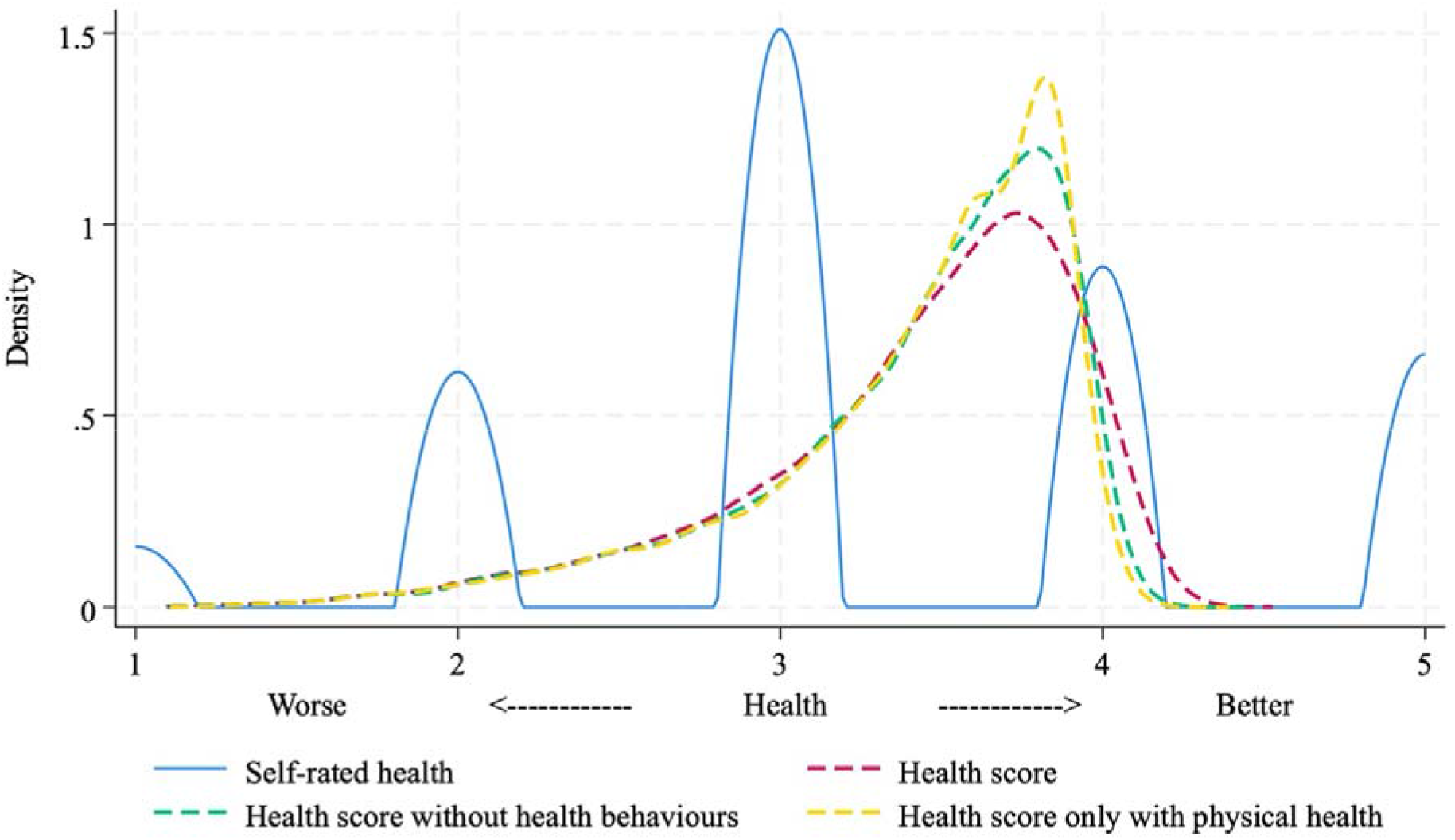

Note: This visualisation is based on the estimation result in Appendix Table A-2.

Appendix Figure A-2. Sampling flow chart and sample size

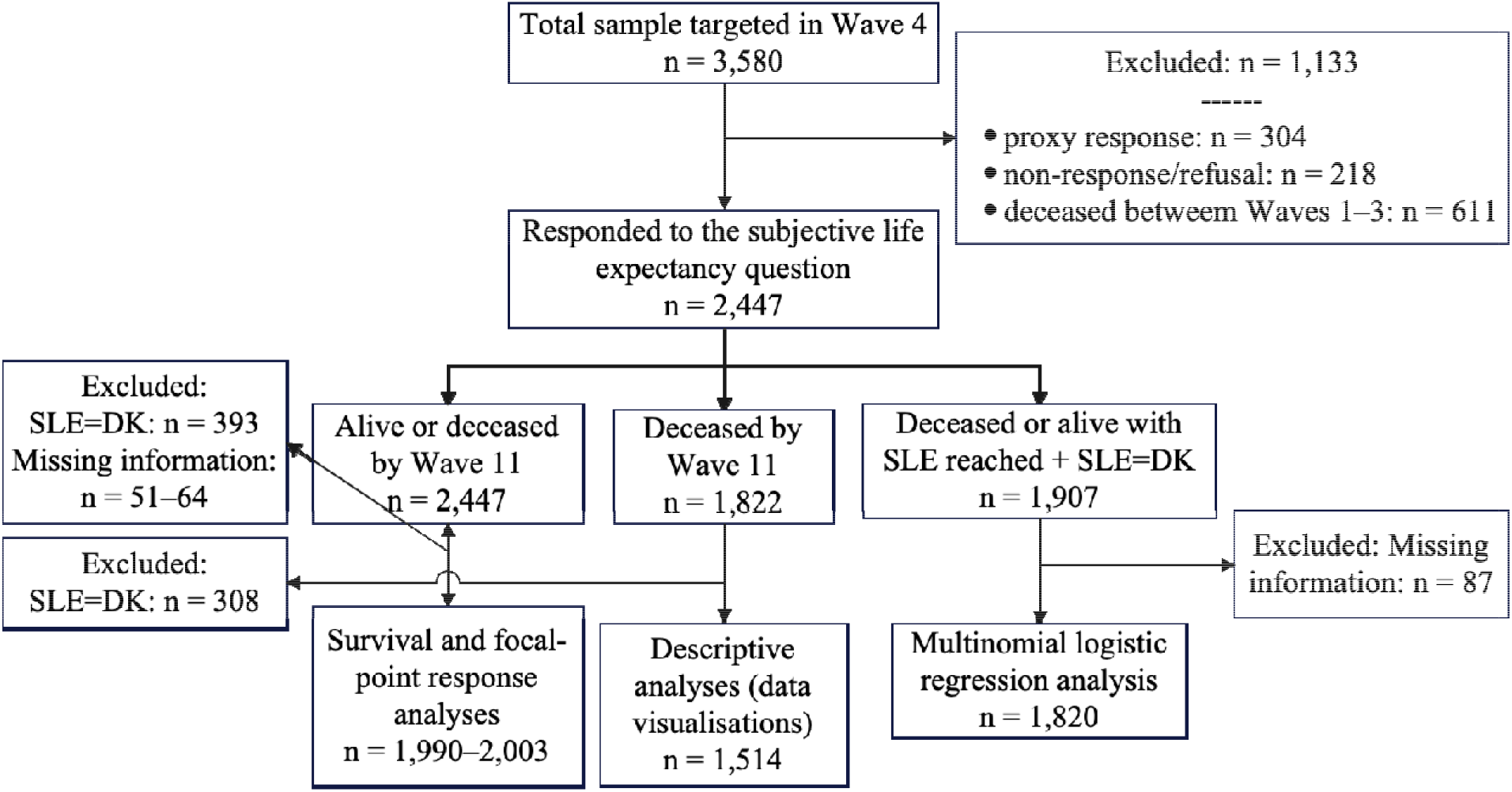

Note: The total targeted sample for Wave 4 consisted of 2,200 individuals from Wave 1, 404 from Wave 2, and 976 newly added individuals; DK represents “do not know” responses to the subjective life expectancy (SLE) question.

Appendix Figure A-3. Respondents’ ages and subjective life expectancy

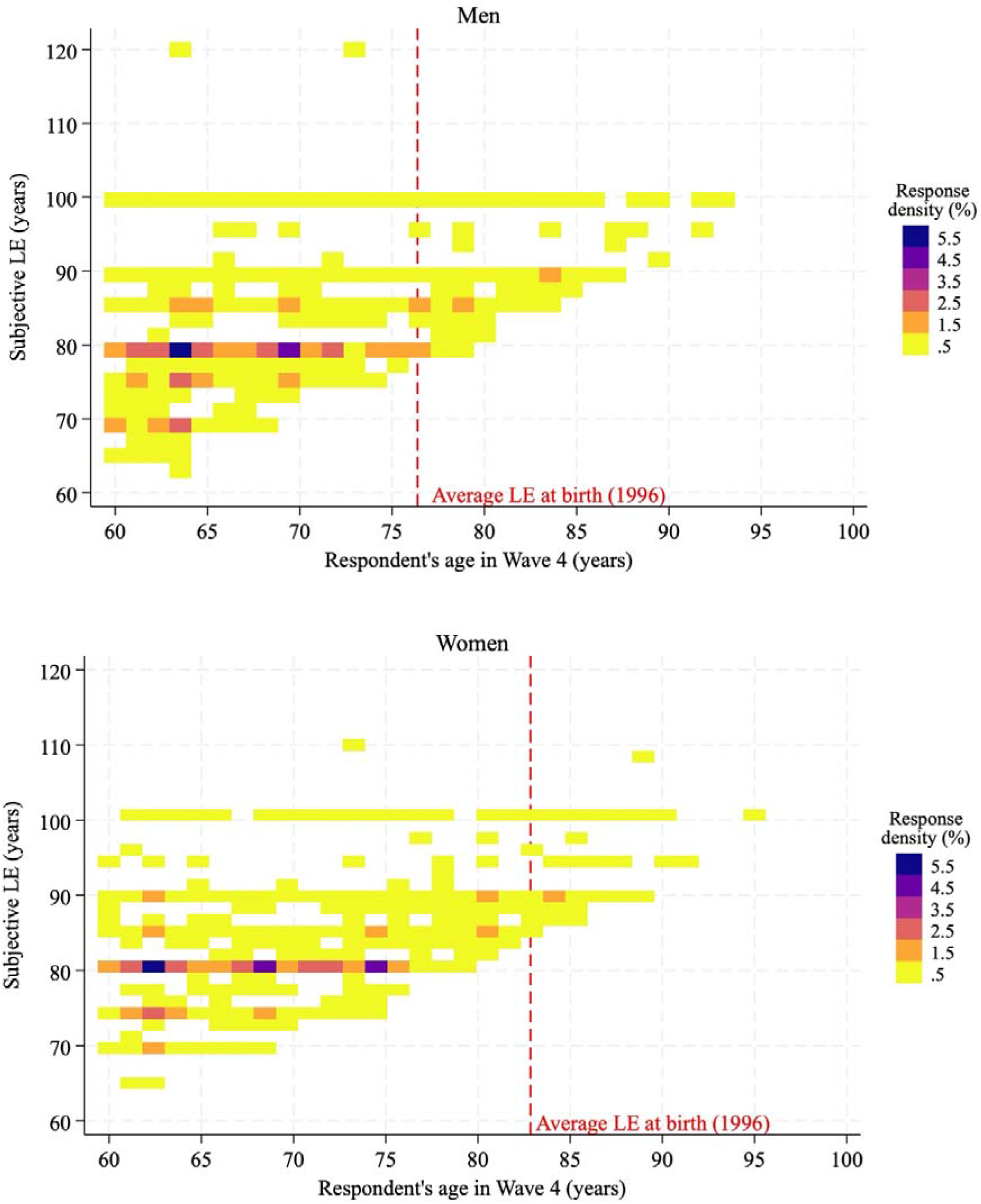

Note: LE stands for life expectancy. This figure was created using the user-written Stata command *heatplot* available on GitHub (https://github.com/benjann/heatplot).

Appendix Figure A-4. Subjective remaining life expectancy and average remaining life expectancy by gender

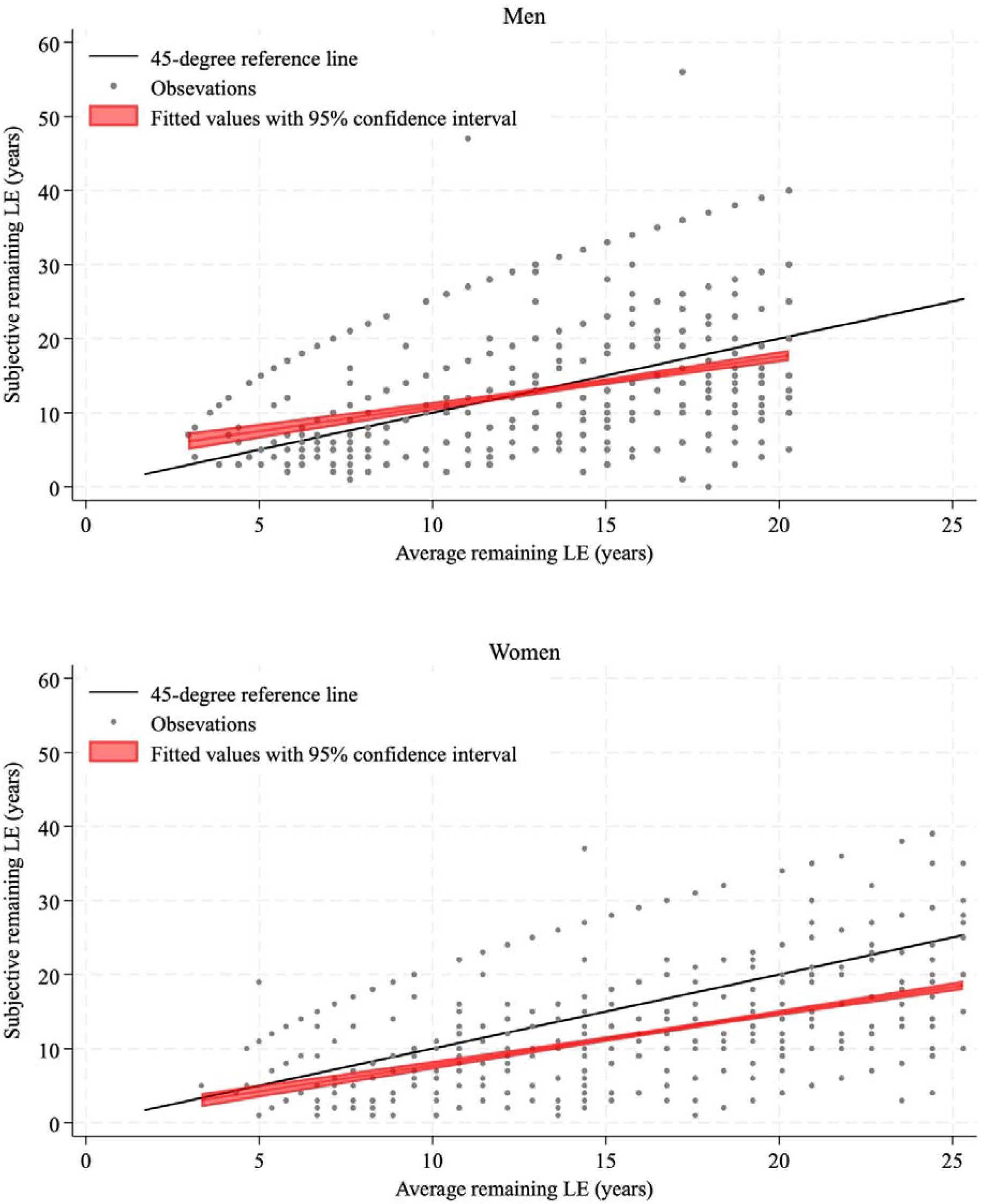

Note: LE stands for life expectancy. This visualisation is obtained for the respondents in Wave 4, with the information on subjective life expectancy available (men: n = 920, women: n = 1,134). Average remaining life expectancy was obtained from official abridged life table estimates and matched to respondents’ ages and genders at Wave 4.

## Data Availability

The data up to wave 9 is available and can be requested to access from the website of the Social Science Japan Data Archive, The University of Tokyo. The data from more recent waves are currently unavailable for external users but will be made available in due course.

https://csrda.iss.u-tokyo.ac.jp/english/

## References

1. Bae, J., Kim, Y.Y., & Lee, J.S. (2017). Factors associated with subjective life expectancy: Comparison with actuarial life expectancy. J Prev Med Public Health, 50, 240–250.

2. Bago d’Uva, T., O’Donnell, O., & van Doorslaer, E. (2020). Who can predict their own demise? Heterogeneity in the accuracy and value of longevity expectations. The Journal of the Economics of Ageing, 17.

3. Balaj, M., Henson, C.A., Aronsson, A., Aravkin, A., Beck, K., Degail, C., et al. (2024). Effects of education on adult mortality: a global systematic review and meta-analysis. The Lancet Public Health, 9, e155–e165.

4. Bell, D.N.F., Comerford, D.A., & Douglas, E. (2020). How do subjective life expectancies compare with mortality tables? Similarities and differences in three national samples. The Journal of the Economics of Ageing, 16.

5. Bound, J., Schoenbaum, M., Stinebrickner, T.R., & Waidmann, T. (1999). The dynamic effects of health on the labor force transitions of older workers. Labour Economics, 6, 179–202.

6. Brouwer, W.B., & van Exel, N.J. (2005). Expectations regarding length and health related quality of life: some empirical findings. Soc Sci Med, 61, 1083–1094.

7. Carstensen, L.L. (2006). The influence of a sense of time on human development. Science, 312, 1913–1915.

8. Coe, N.B., & Zamarro, G. (2011). Retirement effects on health in Europe. Journal of Health Economics, 30, 77–86.

9. Dai, T., Sun, W., & Webb, A. (2023). How do subjective mortality beliefs affect the value of social security and the optimal claiming ages? International Studies of Economics, 19, 92–116.

10. DeSalvo, K.B., Bloser, N., Reynolds, K., He, J., & Muntner, P. (2006). Mortality prediction with a single general self-rated health question. A meta-analysis. Journal of General Internal Medicine, 21, 267–275.

11. Donnelly, R., Umberson, D., & Pudrovska, T. (2020). Family member death and subjective life expectancy among black and white older adults. J Aging Health, 32, 143–153.

12. Dowd, J.B., & Zajacova, A. (2007). Does the predictive power of self-rated health for subsequent mortality risk vary by socioeconomic status in the US? International Journal of Epidemiology, 36, 1214–1221.

13. Elder, T.E. (2013). The predictive validity of subjective mortality expectations: evidence from the Health and Retirement Study. Demography, 50, 569–589.

14. Foltyn, R., & Olsson, J. (2024). Subjective life expectancies, time preference heterogeneity, and wealth inequality. Quantitative Economics, 15, 699–736.

15. Griffin, B., Hesketh, B., & Loh, V. (2012). The influence of subjective life expectancy on retirement transition and planning: A longitudinal study. Journal of Vocational Behavior, 81, 129–137.

16. Griffin, B., Loh, V., & Hesketh, B. (2013). A mental model of factors associated with subjective life expectancy. Soc Sci Med, 82, 79–86.

17. Grossman, M. (2000). The human capital model. In A.J. Culyer, & J.P. Newhouse (Eds.), Handbook of Health Economics pp. 347–408). New York: Elsevier.

18. Hamermesh, D.S. (1985). Expectations, life expectancy, and economic behavior. The Quarterly Journal of Economics, 100.

19. Hurd, M.D., & McGarry, K. (2002). The predictive validity of subjective probabilities of survival. The Economic Journal, 112, 966–985.

20. Hurd, M.D., Smith, J.P., & Zissimopoulos, J.M. (2004). The effects of subjective survival on retirement and Social Security claiming. Journal of Applied Econometrics, 19, 761–775.

21. JAHEAD/NSJE Project Group. (n.d.). Japanese Aging and Health Dynamics Study: National Survey of the Japanese Elderly.

22. Karmarkar, U.R. (2023). Gender differences in "optimistic" information processing in uncertain decisions. Cognitive, Affective & Behavioral Neuroscience, 23, 827–837.

23. Kim, J.H., & Kim, J.M. (2017). Subjective life expectancy is a risk factor for perceived health status and mortality. Health Qual Life Outcomes, 15, 190.

24. Kleinjans, K.J., & Soest, A.V. (2013). Rounding, focal point answers and nonresponse to subjective probability questions. Journal of Applied Econometrics, 29, 567–585.

25. Li, Z., Zhang, Y., Wu, M., & Yang, J. (2024). Is subjective life expectancy stronger in older adults with more physical activity? Evidence from China. *Geriatric Nursing (New York*, N.Y.), 59, 646–652.

26. Lusardi, A., & Mitchell, O.S. (2014). The economic importance of financial literacy: Theory and evidence. Journal of Economic Literature, 52, 5–44.

27. Malanchini, M., Rimfeld, K., Allegrini, A.G., Ritchie, S.J., & Plomin, R. (2020). Cognitive ability and education: How behavioural genetic research has advanced our knowledge and understanding of their association. Neuroscience and Biobehavioral Reviews, 111, 229–245.

28. Manski, C.F. (2004). Measuring expectations. Econometrica, 72, 1329–1376.

29. Ministry of Health, Labor and Welfare. (2019). The 2019 actuarial valuation and the financial implications of the reform options (p.426).

30. Ministry of Health, Labor and Welfare. (2026). Annual Report on the Employees’ Pension Insurance and the National Pension.

31. Ministry of Health, Labour and Welfare. (2025). An overview of an abridged life table 2024.

32. National Institute of Population and Social Security Research. (2023). Population projections for Japan.

33. Niimi, Y., & Horioka, C.Y. (2019). The wealth decumulation behavior of the retired elderly in Japan: The relative importance of precautionary saving and bequest motives. Journal of the Japanese and International Economies, 51, 52–63.

34. Nivalainen, S. (2022). Early pension claiming and expected longevity: A register-based study on the take-up of the partial old-age pension in Finland. *Work*, Aging and Retirement, 8, 264–272.

35. O’Donnell, O., Van Doorslaer, E., Wagstaff, A., & Lindelow, M. (2008). Analyzing health equity using household survey data: a guide to techniques and their implementation. Washington, DC: World Bank Group.

36. OECD. (2025). Pensions at a Glance 2025: OECD and G20 Indicators. Paris: OECD Publishing.

37. Patwardhan, V., Gil, G.F., Arrieta, A., Cagney, J., DeGraw, E., Herbert, M.E., et al. (2024). Differences across the lifespan between females and males in the top 20 causes of disease burden globally: a systematic analysis of the Global Burden of Disease Study 2021. The Lancet Public Health, 9, e282–e294.

38. Post, T., & Hanewald, K. (2013). Longevity risk, subjective survival expectations, and individual saving behavior. Journal of Economic Behavior & Organization, 86, 200–220.

39. Rappange, D.R., Brouwer, W.B., & van Exel, J. (2016a). A long life in good health: subjective expectations regarding length and future health-related quality of life. Eur J Health Econ, 17, 577–589.

40. Rappange, D.R., Brouwer, W.B., & van Exel, J. (2016b). Rational expectations? An explorative study of subjective survival probabilities and lifestyle across Europe. Health Expectations, 19, 121–137.

41. Rappange, D.R., van Exel, J., & Brouwer, W.B. (2017). A short note on measuring subjective life expectancy: survival probabilities versus point estimates. Eur J Health Econ, 18, 7–12.

42. Ruud, P.A., Schunk, D., & Winter, J.K. (2025). Uncertainty causes rounding: an experimental study. Experimental Economics, 17, 391–413.

43. Seaman, S.R., & White, I.R. (2013). Review of inverse probability weighting for dealing with missing data. Statistical Methods in Medical Research, 22, 278–295.

44. Smith, V.K., Taylor, D.H., & Sloan, F.A. (2001). Longevity expectations and death: Can people predict their own demise? The American Economic Review, 91, 1126–1134.

45. Sticha, A., & Sekita, S. (2023). The importance of financial literacy: Evidence from Japan. Journal of Financial Literacy and Wellbeing, 1, 244–262.

46. Thaler, Richard H., & Benartzi, S. (2004). Save More Tomorrow™: Using Behavioral Economics to Increase Employee Saving. Journal of Political Economy, 112, S164–S187.

47. van Solinge, H., & Henkens, K. (2010). Living longer, working longer? The impact of subjective life expectancy on retirement intentions and behaviour. European Journal of Public Health, 20, 47–51.

48. van Solinge, H., & Henkens, K. (2018). Subjective life expectancy and actual mortality: results of a 10-year panel study among older workers. Eur J Ageing, 15, 155–164.

49. Wagstaff, A. (2005). The bounds of the concentration index when the variable of interest is binary, with an application to immunization inequality. Health Economics, 14, 429–432.

50. Yang, Q., Ye, Z., & Chen, R. (2024). Working longer or working harder? Subjective survival expectations and labor supply in China. International Review of Economics & Finance, 91, 827–847.

